# Bridging Computational and Clinical Strategies to Improve Presurgical Identification of Epileptogenic Networks

**DOI:** 10.1101/2025.08.22.25334229

**Authors:** Tena Dubcek, Kristina Koenig, Adham Elshahabi, Debora Ledergerber, Rafael Polania, Lukas Imbach

**Affiliations:** ETH Zurich, Department of Health Sciences and Technology, Switzerland; Swiss Epilepsy Center, Klinik Lengg, Zurich, Switzerland; Neuroscience Center Zurich, University of Zurich and ETH Zurich, Switzerland

## Abstract

About one third of epilepsy patients are drug-resistant. Resective surgery remains a key treatment option but depends critically on accurate identification of the seizure onset zone (SOZ), which is still guided mainly by subjective visual inspection of electrophysiological signals. Network-based metrics derived from intracranial EEG (iEEG) have recently shown promise for SOZ identification, but their interpretation has remained disconnected from standard clinical procedures, while the reported performance often hinges on the choice of machine learning classifiers and summary scores.

We analyzed interictal stereotactic EEG (sEEG) recordings from 20 patients undergoing presurgical evaluation, including cortical and subcortical implantations, with clinical mapping via electrical stimulation. We constructed patient-specific dynamic network models and compared the values of four corresponding metrics of network vulnerability (outgoing fragility, incoming fragility, source influence, sink connectivity) that were previously proposed as promising SOZ markers, with stimulation-evoked discharges. We also simulated virtual thermocoagulation by removing the clinically coagulated nodes and testing whether the resulting network changes went beyond pure network size reduction.

The network metrics correlated with epileptiform discharges evoked by 50 Hz intracranial stimulation, directly linking model-based fragility with interictal epileptiform discharges evoked in clinical stimulation mapping. Using virtual thermocoagulation, we showed that the network models can predict the consequences of lesioning, capturing both local and global effects depending on individual network architecture. Across patients, the network metrics consistently distinguished SOZ from non-SOZ contacts and yielded stable conclusions across time, conditions and perturbation properties, supporting their reliability.

Together, these findings show that iEEG-based network models provide clinically meaningful and interpretable markers of brain responsiveness to electrical stimulation, and can be used to predict the consequences of virtual resections. By relying only on interictal recordings, they avoid the clinical and technical challenges of capturing seizures and instead offer a personalized framework that complements presurgical mapping and guides surgical planning in drug-resistant epilepsy.

## Introduction

Epilepsy is a common neurological disorder, affecting approximately 1% of the global population (Devinsky *et al*., 2018). About one third of epilepsy patients are drug-resistant, meaning they do not respond to two or more anticonvulsive medications (Devinsky *et al*., 2018). Motivated by these challenging cases, alternative therapeutic approaches have been developed, including resective epilepsy surgery (Ryvlin, Cross and Rheims, 2014; Zijlmans, Zweiphenning and van Klink, 2019; Cross *et al*., 2022) and brain stimulation (Ryvlin *et al*., 2021; Xue *et al*., 2022). These therapies heavily depend on understanding the epileptic network—a distributed set of brain regions whose abnormal and coordinated activity underlies seizure generation and propagation (Kramer and Cash, 2012; Li *et al*., 2021). A critical step for successful therapy is accurately identifying the resection or stimulation targets that can effectively modulate the pathological network state and achieve seizure freedom (Chowdhury *et al*., 2021). In clinical practice, this is commonly guided by visual inspection of intracranial EEG (iEEG) to define the seizure onset zone (SOZ) (Jayakar *et al*., 2016), the region where seizures are first observed (Chowdhury *et al*., 2021). In addition, interictal epileptiform discharges (Kramer and Cash, 2012; Fitzgerald *et al*., 2021; Thomas *et al*., 2023), high-frequency oscillations (Zijlmans *et al*., 2012; Frauscher *et al*., 2017; Nevalainen *et al*., 2020; Remakanthakurup Sindhu, Staba and Lopour, 2020; Tamilia *et al*., 2021; Cserpan *et al*., 2022) (HFOs) and other focal EEG biomarkers are used as biomarkers to support localization. However, all these features represent local properties of the signal, while it is increasingly recognized that epilepsy is a network disorder involving interactions across multiple brain regions (Kramer and Cash, 2012). Ideally, clinicians would have access to objective and personalized metrics and models capable of capturing these network dynamics and identifying the network regions most responsible for seizure generation and propagation. Several quantitative and modeling approaches have recently emerged to address this need (Goodfellow *et al*., 2016; Jirsa *et al*., 2017; An *et al*., 2019; Larivière *et al*., 2021; Bernabei *et al*., 2022; Cao *et al*., 2022; Kerr and McFarlane, 2023; Runfola *et al*., 2023; Sinha *et al*., 2023; Lin *et al*., 2024; Wang *et al*., 2024). Nevertheless, a common challenge is their interpretability within existing clinical frameworks including electrical stimulation based mapping (Trébuchon and Chauvel, 2016), as well as their feasibility in clinical practice, both necessary for clinician acceptance and practical use in epileptology.

An outstanding example illustrating this issue is the recently introduced metric of neural fragility (Li *et al*., 2021; Myers *et al*., 2025), a model-based quantitative index which was retrospectively demonstrated to hold significant potential for guiding surgical planning in drug-resistant epilepsy. Neural fragility involves the construction of personalized iEEG-based dynamical models of the (ictal) epileptic network (Li *et al*., 2021; Dubcek *et al*., 2025), providing model-based insights into how easily perturbations of each network node can destabilize the network and trigger the seizure onset (Li *et al*., 2021). Despite its promise, neural fragility has not yet seen broad clinical adoption, due to both practical and conceptual challenges. The method relies on relatively advanced principles from nonlinear dynamics, which may hinder accessibility and integration into clinical workflows. While it shows strong potential for identifying the seizure onset zone (SOZ) (Li *et al*., 2021), its interpretation and alignment with established clinical practices (Trébuchon and Chauvel, 2016; Ritaccio, Brunner and Schalk, 2018; Grande, Ihnen and Arya, 2020; Bernabei *et al*., 2023) remain active areas of investigation. Other concerns regarding the generalizability and robustness of neural fragility include its sensitivity to different types of perturbations, the source data selection (particularly how effectively models derived from interictal recordings perform), and the potential inclusion of subcortical structures. Moreover, other proposed metrics sharing similar rationales (Gunnarsdottir *et al*., 2022) have not been adequately differentiated or discussed comparatively.

In this study, we first explicitly addressed these limitations using a cohort of 20 epilepsy patients implanted with stereotactic depth electrodes (sEEG). Based on interictal recordings, we constructed dynamic network models (Li *et al*., 2021; Dubcek *et al*., 2025) and evaluated the following four network-based metrics for each channel: outgoing fragility, incoming fragility, source influence, and sink connectivity (Li *et al*., 2021; Gunnarsdottir *et al*., 2022; Bernabei *et al*., 2023). We demonstrated that all four metrics are closely related, reliably differentiate SOZ from non-SOZ channels, and remain stable throughout time and perturbation properties. Motivated by the fact that presurgical evaluations of epilepsy often involve iEEG-based brain stimulation mapping (Trébuchon and Chauvel, 2016), which essentially serves as an experimental counterpart to neural fragility, we analyzed how the level of epileptiform discharges evoked by 50 Hz intracranial direct current stimulation relates to the network-based metrics. In addition, we showed how network-based models could be used for virtual thermocoagulation and prediction of resection consequences.

## Methods

### Patient population and electrode implantation

We included 20 patients with pharmacoresistant focal epilepsy (9 females, mean age 33±12 years) who underwent intracranial EEG (iEEG) recordings with stereo electroencephalography (sEEG) as part of their presurgical evaluation at the Swiss Epilepsy Center (Fig. 1A). All patients were candidates for epilepsy surgery. The sEEG procedure (including reduction of medication, direct electrical stimulation, and duration of recording) was performed based solely on clinical decisions.

**Figure 1.**
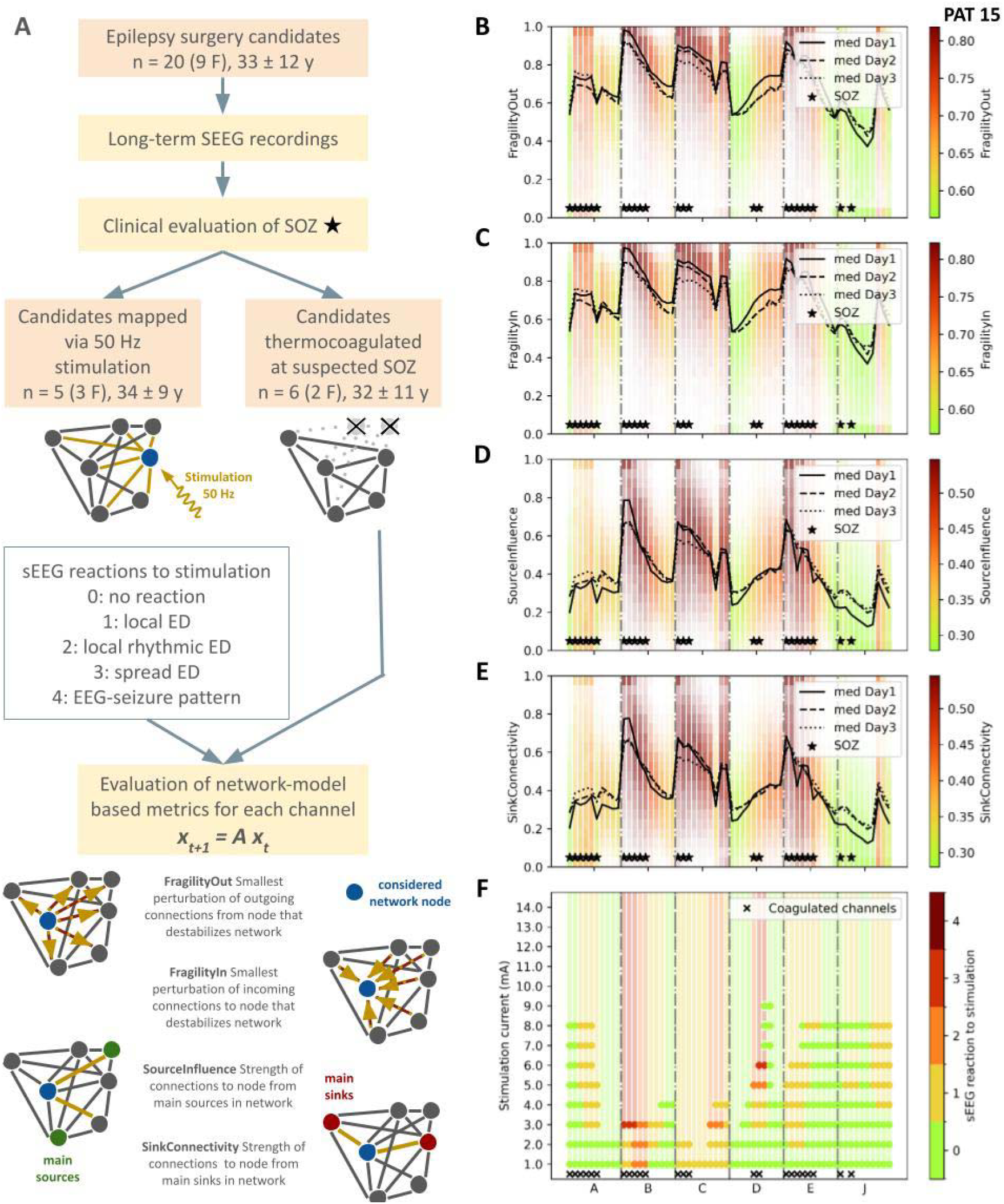
Overview — **A** Schematic of computational and clinical pipeline. **B–E** Dynamic network-based metrics for one exemplary patient. black lines indicate daily medians. Shaded distributions show channel-wise values per day, colors reflect daily medians for visual comparison. Black stars mark clinically defined SOZ channels. Electrode names are shown at the bottom; each electrode contains multiple contacts. **B** Outgoing fragility. **C** Incoming fragility. **D** Source influence. **E** Sink connectivity. **F** Quantified reactions to 50 Hz stimulation in same patient. Filled circles denote tested electrode–current pairs. Background shading denotes maximal observed reaction per electrode across all tested currents. Black crosses mark thermally coagulated channels.

The implantation strategy was individualized for each patient and based on prior noninvasive evaluations. Electrodes from DIXI-medical Microdeep® and AD-TECH-medical Spencer were stereotactically implanted in cortical and subcortical brain regions suspected to be involved in seizure generation. Electrode locations were verified using preimplantation and postimplantation magnetic resonance imaging (MRI). A detailed overview of implanted electrodes, antiseizure medications, and clinical profiles of all patients is found in Tab. 1.

**Table 1.** Patient demographics: age range, sex, antiseizure medications during iEEG recording, whether mapping by electrical stimulation was performed, whether thermocoagulation was performed, and whether resection was performed and with which outcome (Engel class). — BRV = Brivaracetam; CBD = Cannabidiol; CEN = Cenobamate; CLB = Clobazam; CLN = Clonazepam; CLZ = Clonazepam; ESL = Eslicarbazepine acetate; LCM = Lacosamide; LEV = Levetiracetam; LTG = Lamotrigine; OXC = Oxcarbazepine; PER = Perampanel; PGB = Pregabalin; PHE = Phenytoin; PHT = Phenytoin; VPA = Valproate; ZNS = Zonisamide.

| PAT | Age Range | Sex | ASM at iEEG | Electr. Stim. | Therm. Coag. | Resect. | Resect. Outcome |
| --- | --- | --- | --- | --- | --- | --- | --- |
| 1 | 26-30 | M | LEV, LCM, CLB, PER | N | N | N | - |
| 2 | 26-30 | F | BRV, CLN | N | N | N | - |
| 3 | 26-30 | F | LEV, PER | N | N | Y | IA |
| 4 | 56-60 | M | BRV, LCM | N | N | N | - |
| 5 | 26-30 | M | LEV, LTG, PER | N | N | Y | IA |
| 6 | 26-30 | M | LTG, BRV | N | N | N | - |
| 7 | 31-35 | M | ZNS | N | N | Y | IIB |
| 8 | 16-20 | M | BRV, LTG | N | N | Y | IA |
| 9 | 16-20 | M | LEV, LCM | N | N | N | - |
| 10 | 06-10 | F | OXC | Y | N | Y | III |
| 11 | 16-20 | M | LEV, OXC | Y | Y | Y | IA |
| 12 | 41-45 | M | LTG, VPA, LCM | Y | Y | N | - |
| 13 | 11-15 | F | PER, OXC | N | N | Y | IB |
| 14 | 36-40 | F | CEN | N | N | N | - |
| 15 | 21-25 | F | PHE, BRV, ESL, ZNS, CLB | Y | Y | Y | III |
| 16 | 31-35 | F | PER, LEV, LTG | N | N | N | - |
| 17 | 41-45 | M | ESL, PER | Y | Y | N | - |
| 18 | 26-30 | M | LTG | Y | Y | N | - |
| 19 | 31-35 | F | BRV, CBD, LCM | Y | Y | N | - |
| 20 | 41-40 | F | LEV, PHT, PGB, CLZ | Y | N | N | - |

The study was approved by the local ethical committee (Kantonale Ethikkommission Zürich, Approval PB 2016–02055), and all patients or their legal representatives provided written informed consent in accordance with the Declaration of Helsinki. However, this consent did not include a provision for making individual data freely accessible.

### Clinical mapping and radiofrequency thermocoagulation

A subgroup of five candidates underwent 50 Hz stimulation via electrodes from Micromed, to map brain activity and identify functional eloquent brain regions (Fig. 1A). The sEEG reactions (changes in baseline EEG) were quantified by two independent clinicians, using the following scale: (0) no reaction, (1) local epileptiform discharge, (2) local rhythmic epileptiform discharges, (3) spread epileptiform discharges, (4) EEG-seizure pattern.

During the presurgical evaluation of all candidates, the responsible clinicians defined the suspected seizure onset zone (SOZ) based on the combined analysis of sEEG and scalp EEG during the recorded seizures. Based on this evaluation, each sEEG channel was classified as either SOZ or non-SOZ and treated accordingly in all subsequent analyses.

A subgroup of 6 candidates underwent local brain lesioning by radiofrequency thermocoagulation (coagulation device from Inomed Medizintechnik GmbH) around electrode contacts that were identified as belonging to SOZ, but not part of an eloquent area (Pichardo-Rojas *et al*., 2025) (Fig. 1**A**).

### Dynamic network models of sEEG

We modeled the interictal brain dynamics by a discrete-time linear approximation of the form

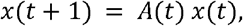

Where *x* (*t*) ∈ R*^n^* is the multichannel sEEG signal at time *t*, and *A* ∈ *R*^*n*×*n*^ is the state transition matrix capturing effective directed interactions between channels. The matrix *A* was estimated independently over sliding windows of 0.5 s duration and with 0.5 s shift, sampled down to 200 Hz, as frequencies higher than 100 were not considered significant for the analysis. In each window, *A* was obtained by solving a regression model with l2-regularization (α*Ridge* = 10^-5^), yielding a time-varying sequence of linear models that formed the basis of all subsequent metrics.

Multiple 10-minute long sEEG recordings were analyzed for each patient. In contrast to previous studies focused on ictal activity, our analysis was conducted entirely on interictal recordings, which offer more stable conditions, better comparability across patients, and avoid the confounding effects of seizure-related dynamics. For all patients, data recorded in the morning, afternoon, and early night two days after implantation were used for analyzing temporal robustness of the network-based metrics. For patients who underwent stimulation mapping, data recorded at approximately the same time in the three consecutive days prior to stimulation was used. For patients who underwent radiofrequency thermocoagulation, the mean of morning, afternoon, and night metric values was used in model-based virtual thermocoagulation.

### Outgoing and incoming fragility

To quantify the susceptibility of each node to destabilizing the network, we computed neural fragility based on structured perturbation analysis (Li *et al*., 2021). Intuitively, fragility reflects how close a given brain region is to tipping the whole network into a seizure-prone state: fragile nodes are those where even small changes in their connections can destabilize the system, making them likely contributors to seizure initiation.

For each estimated matrix *A*(*t*), frequency dependent inverse fragility is defined as the minimal norm of a perturbation to the k^th^ row or column of *A* that pushes one of its eigenvalues *λ* to the unit circle,

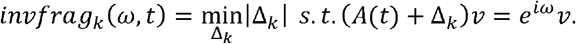

Column perturbations (Δ_*k*_ = *e*_*k*_*⌈*^*T*^) simulate changes to incoming connections, while row perturbations 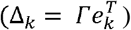 affect outgoing projections. The constrained minimization can be rewritten as

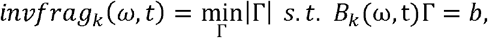

with, for column perturbations,

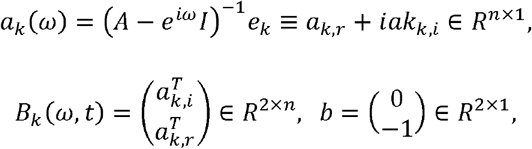

and for row perturbations,

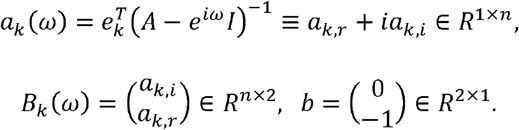

The closed-form solution, obtained using the Lagrange multipliers method, is

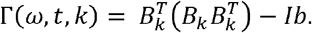

Node fragility is then obtained from

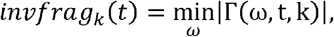

with *ω* ∈ [0.5,100] Hz, by inversely linearly normalizing *invfrag*_*k*_ across all nodes *k* within each time window *t*,

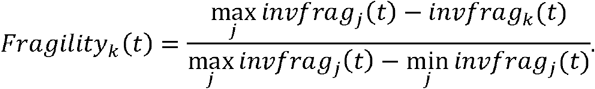

We computed normalized fragility separately for column (outgoing) and row (incoming) perturbations, resulting in two complementary fragility maps per time window, 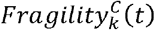 and 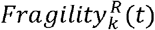. Since outgoing and incoming fragility proved extremely highly correlated (see Results), we also defined a single fragility metric *Fragility*_*k*_(*t*) as their average. To obtain time-aggregated fragility profiles, the normalized fragility values were summarized across windows by taking the median over time.

### Source influence and sink connectivity

To complement fragility, we also extracted source influence and sink connectivity metrics (Gunnarsdottir *et al*., 2022) from the dynamic network models. Intuitively, the source–sink framework characterizes the directional flow of influence in the network: sources are nodes that strongly drive others, while sinks are strongly driven. In epilepsy, seizure onset zones seem to often appear as abnormal sinks, reflecting their heightened recruitment by surrounding regions.

Based on the estimated matrix e computed the total absolute incoming and outgoing weights for each node *k*

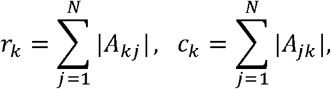

and their rank-normalized values *rr*_*k*_ and *cr*_*k*_ across all nodes. The sink index was the proximity to the N ideal sink point 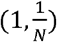 in rank space

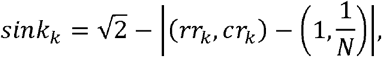

and the source index analogously to 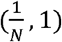 These indices were then used to compute higher-order connectivity features, source influence,

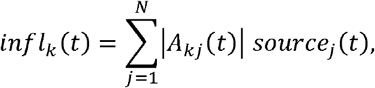

and sink connectivity,

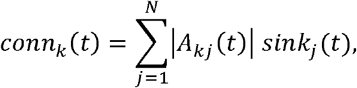

and their normalized versions

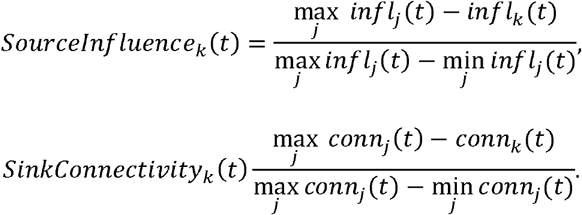

Since these two measures later proved to be highly correlated (see Results), we also defined a single metric as their average,

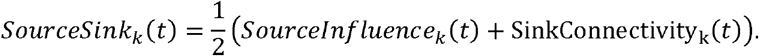

Time aggregation was done analogously as for fragility.

### Statistical methods

Clinical data is rarely Gaussian due to relatively small sample sizes and inter-patient variability. We therefore used nonparametric, permutation-based analyses for all statistical comparisons. In particular, permutation tests were used to assess correlations between network metrics by permuting channel-wise metric values within each patient. For group comparisons, such as between SOZ and non-SOZ channels, between reacting and non-reacting channels, or between different times of day, group labels were permuted while preserving the underlying metric values. In all cases, the observed statistic was compared to the empirical null distribution generated from 10000 permutations. (*p* < 0.001). The significance threshold was set at α= 0.05, and Bonferroni correction was applied For visualization, statistical significance is denoted by asterisks: ^*^ (*p* < 0.05), ^**^ (*p* < 0.01), and ^***^ when performing multiple comparisons within individual patients (e.g., for pairwise metric correlations or daytime condition effects).

In analyses involving thermocoagulation, we computed a null distribution of network metric changes using repeated random removal of channels. The null distributions obtained for each channel were then bootstrapped with 5000 repetitions, and their 97.5th percentile was used to estimate whether changes after removal of clinically coagulated nodes exceeded the chance and effects that come exclusively from a change in network size. This empirical approach provided a patient-specific control for interpreting network effects.

## Results

The sEEG recordings of each patient were used for data-driven extraction of the network dynamics (encoded in the dynamic connectivity matrices *A*) and the evaluation of the corresponding network-based metrics for each sEEG channel (Fig. 1**A**). Outgoing and incoming fragility was defined as the normalized time-median of the smallest perturbation to outgoing and incoming connections of a node that destabilizes the whole network Fig. 1**BC**). Source influence reflected the normalized time-median of the strengths of connections from the main sources in the network (Fig. 1**D**), while sink connectivity reflected the normalized time-median of the strengths of connections from the main sinks in the network (Fig. 1**E**). These network-derived measures were then compared with the severity of epileptiform discharges evoked during clinical 50 Hz intracranial stimulation mapping (Fig. 1**F**). The detailed results for all other patients can be found in Supplementary Information.

### Network-based metrics point toward SOZ and remain stable despite EEG variability

We first assessed the ability of these network-based metrics to point toward the SOZ based on exclusively interictal data of all patients, having implanted both cortical and subcortical electrodes. In a personalized manner, we compared the distributions of time-aggregated fragility and source-sink values (see Methods for definitions of *Fragility* and *SourceSink*) between EEG channels inside and outside the SOZ for each patient (Fig. 2**AB** left). The metrics evaluated from sEEG data recorded in the morning showed significant differences in distribution means inside and outside the SOZ for 12 of the 20 patients. Interestingly, patients who showed significant differences in fragility distribution also showed significant differences in source-sink distributions, although the significance level could vary. By comparing the distributions of patient-wise means inside and outside the SOZ (Fig. 2**AB** non-SOZ right), we could show that fragility and source-sink values are significantly higher in SOZ than in regions on a group level: fragility Δ_*F*_ = 0.09, *p*_*F*_ = 0.017, and source-sink Δ_*ss*_=0.12, *p*_*ss*_ = 0.003

**Figure 2.**
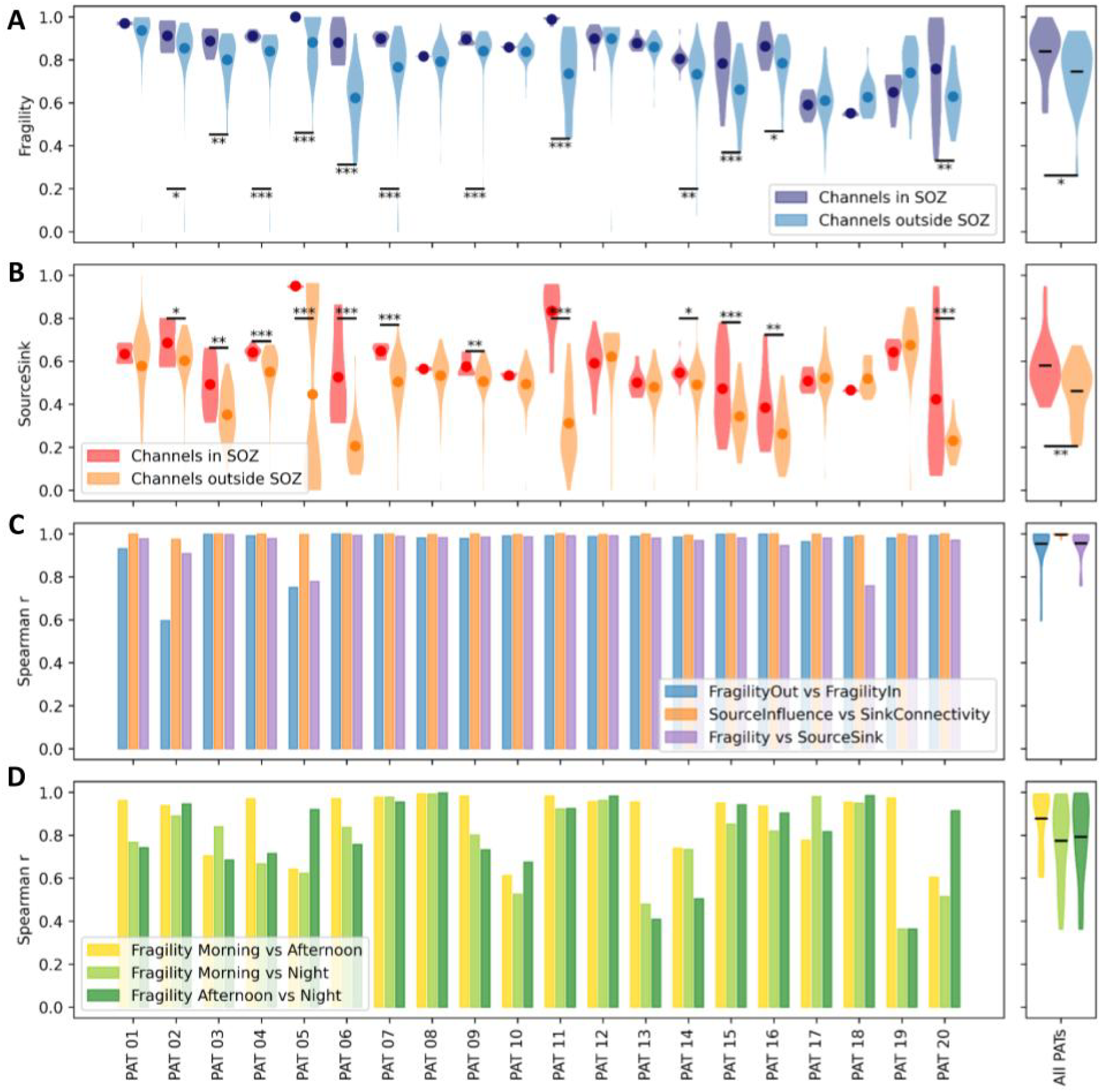
Network-based metrics point toward the SOZ and are extremely robust. — **A** Left: Patient-wise comparison of fragility distributions (average of outgoing and incoming fragility) between channels inside and outside the seizure onset zone (SOZ). Stars indicate levels of statistically significant differences in means based on permutation testing. Right: Distribution of patient-level means for channels inside and outside the SOZ. Black horizontal lines denote the population means. **B** Same as A, but for the source-sink metric (average of source influence and sink connectivity). **C** Left: Patient-wise Spearman correlations between outgoing and incoming fragility, source influence and sink connectivity, and the means of fragility and source-sink metrics. Right: Population distributions and means of the correlation values. **D** Left: Spearman correlations of fragility metrics computed from recordings at different times of day. Right: Population distributions and means of the correlation values.

Next, we aimed to clarify the robustness of the conclusions derived from different network-based metrics. We analyzed the differences in channel dependences of outgoing, incoming fragility, source influence and sink connectivity for each patient. We observed statistically significant correlations for all patients and all metric type pairs (Fig. 2**C** left). On the population level, the Spearman correlations showed the following confidence intervals: outgoing vs incoming fragility *r* ∈ [0.92,0.99], source influence vs sink connectivity *r* ∈ [0.96,1.00], and fragility vs source-sink *r* ∈ [0.70,0.88] (Fig. 2**C** right). Thus, despite their different definitions, all four metrics converge on highlighting the inter-regional dynamics and regions driving network instability.

Finally, we assessed sensitivity to the choice of data time span by comparing results derived from morning, afternoon, and night sEEG recordings (Fig. 2**D** left). We found that the distributions of hannel fragility remained significantly correlated across time for all patients. On the population level, the Spearman correlations showed the following confidence intervals: morning vs afternoon *r* ∈ [0.79,0.97], morning vs night *r* ∈ [0.68,0.86], and afternoon vs night *r* ∈ [0.70,0.88] (Fig. 2**D** right). We further showed that the choice of eigenvalue frequency used to quantify node fragility had only a negligible impact (relative difference Δ < 1% across all channels and all patients for frequencies in range 0.5 − 100 s). Together, these findings indicated that the identified vulnerable regions remain highly stable across time, conditions, and frequency choices, underscoring the robustness and reliability of the approach.

### Model-based fragility correlates with sEEG reactions to stimulation

We used direct electrical stimulation at 50 Hz in a subset of patients during presurgical evaluations to map eloquent brain areas and identify the SOZ. EEG reactions to stimulation quantify how the latter affects the departure from the steady state of individual brain dynamics. This approach offers a way to experimentally probe the network’s reactivity, similar in spirit to model-based perturbation analysis, where simulated perturbations are used to assess how close a given network node is to destabilizing the system. Both approaches aim to reveal the parts of the network that are most capable of driving pathological activity. We therefore expected channels that respond strongly to stimulation to also show high fragility or influence in the model-based metrics.

For each patient, we correlated the values of the fragility and source-sink metrics with the corresponding stimulation-assessed reactivities, and we compared the metric means between reacting and non-reacting channels (Fig. 3). Four of the five stimulated patients showed positive correlations between model-based metric values and the strength of stimulation-evoked responses during presurgical evaluations, with statistical significance varying across patients and metric types (see Fig. 3**AB** for individual Spearman *r* values). The difference in mean network metric values between statistical significance that varied depending on the patient and metric (see Fig. 3**CD** for individual Δ channels that reacted to stimulation and those that did not was positive for all patients, again with and p values).

**Figure 3.**
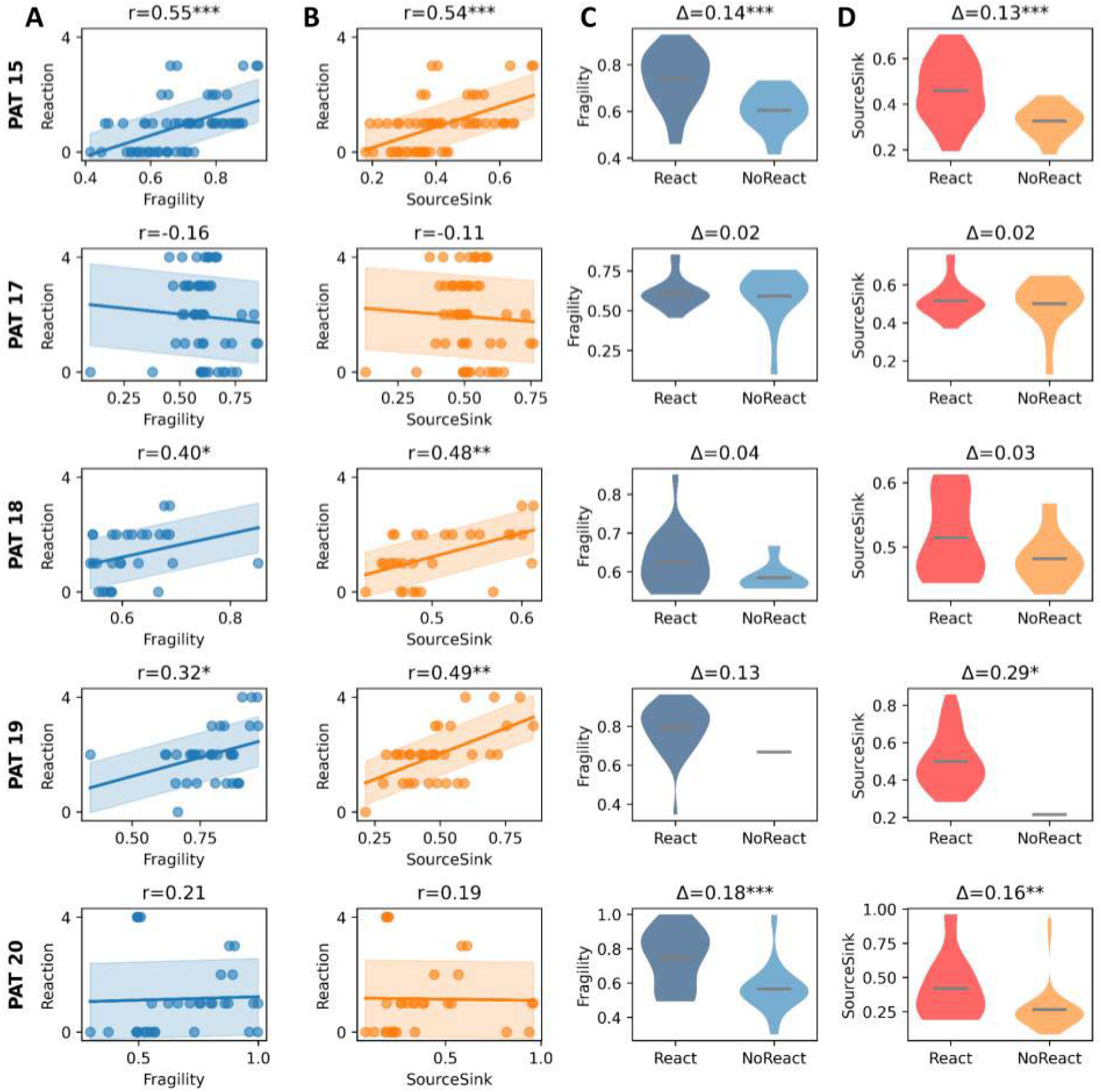
Model-based fragility and source-sink metrics correlate with stimulation-evoked sEEG responses in individual patients. — **A** Correlation of model-based fragility values and level of sEEG reaction to 50 Hz direct electrical stimulation, used to map eloquent brain areas and identify SOZ in presurgical evaluation. The Spearman’s correlation values and statistical significance based on permutation testing are shown at the top of each subplot. **B** Same as A, but for source-sink metric. **C** Distributions and means (vertical lines) of model-based fragility values in sEEG channels that reacted to stimulation and those that did not. The difference in means and the corresponding statistical significance based on permutation testing are shown at the top of each subplot. **D** Same as C, but for source-sink metric.

### Predicting consequences of virtual brain resection via network models

Given that radiofrequency thermocoagulation and surgical interventions target specific brain regions suspected to be responsible for initiating seizures, we asked whether our models and model based metrics could help predict how removing these regions would impact overall network stability. From a modeling perspective, such regions may correspond to nodes whose perturbation induces pathological transitions toward unstable brain states. When performing thermocoagulation or brain resection, the changes in the brain occur locally, but what ultimately matters is how these changes propagate through the broader network. Focusing solely on the SOZ may therefore be shortsighted, as it does not capture downstream or network-wide consequences that may be critical for seizure generation or control. Network models may help uncover these non-local effects.

To address this, we propose an approach for using personalized network models to post-hoc simulate the effects of node removal via thermocoagulation. For each patient, we evaluated how fragile the nodes were when the full network was intact, and how this changed after virtual removal of the nodes that had been clinically coagulated (Fig. 4). To distinguish changes caused by the removal of nodes driving network instability from those caused merely by a reduction in network size, we computed a null distribution by repeatedly removing the same number of randomly selected nodes. This allowed us to determine, for each patient, the network-driven consequences of the local changes that were chosen clinically. We observed patients in whom the entire network became significantly more resilient and less fragile after the virtual removal of a few key nodes (mostly green in Fig. 4), but also patients in whom the virtual removal of the coagulated nodes had almost no significant consequences for the remaining network. Interestingly, in patients whose network-driven changes were relatively weak, we could still observe local effects near the coagulated channels (green changes adjacent to coagulated contacts in Fig. 4). This can be seen as an internal validation, showing that local effects are expected because strong nearby connections, while global changes arise only when permitted by the patient’s network architecture.

**Figure 4.**
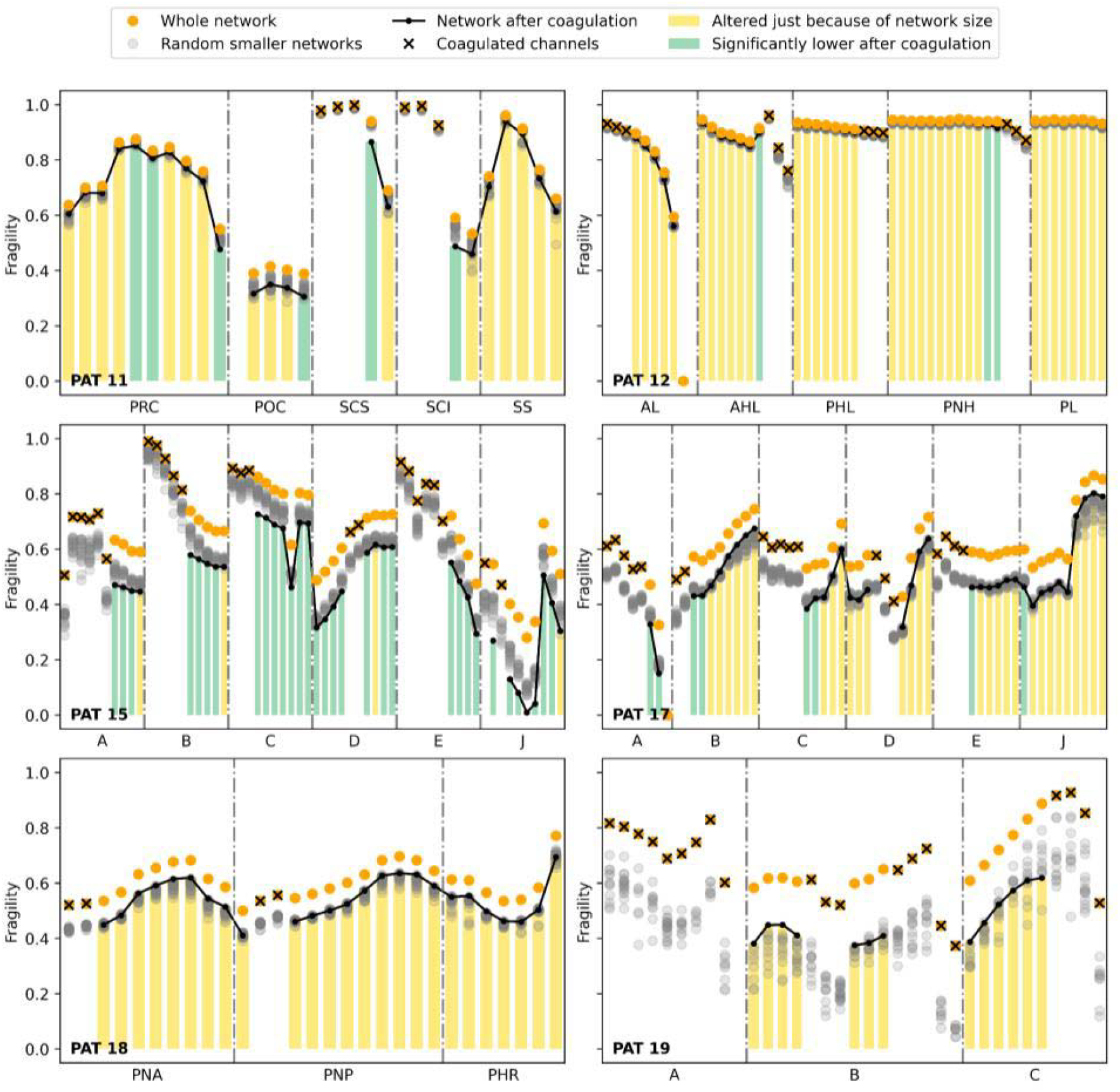
Predicting network-level consequences of brain thermocoagulation using personalized virtual resection models. — Each subplot corresponds to one patient who underwent thermocoagulation of selected electrode contacts. Orange markers denote fragility values for whole, intact network. Black lines denote fragility values after virtual removal of clinically coagulated nodes, signaled with black crosses. Gray transparent markers show the null distribution of fragility values obtained after repeatedly removing the same number of randomly selected nodes. The comparison with the 97.5th percentile of the corresponding bootstrapped distribution is then used to determine the specificity of the observed changes in fragility for the virtual removal of coagulated nodes: Channels with significantly decreased fragility after virtual thermocoagulation are colored in green, the rest in yellow.

### Discussion

In this study, we systematically evaluated a set of network-based metrics, derived from interictal sEEG recordings, to characterize the stability and responsiveness of the epileptic brain network in focal drug-resistant epilepsy. Going beyond traditional biomarker development, we focused on bridging computational modeling with clinical application. We demonstrated that these interictal metrics correlate with sEEG responses evoked by intracranial stimulation and can predict the effects of virtual thermocoagulation, thereby offering a personalized, model-based perspective on stimulation and resection strategies. To support their interpretability and reliability, we first established that the four metrics, although conceptually distinct, are highly correlated, discriminate SOZ from nonSOZ regions, and remain remarkably stable across times of day and perturbation properties. This coherence suggests they capture a common, high-level property of network dynamics, likely linked to the non-linear functional connectivity and global stability landscape of the epileptic brain (Kramer and Cash, 2012).

Importantly, our findings were obtained from interictal recordings alone, avoiding the clinical and technical challenges associated with capturing ictal activity. In contrast to conventional biomarkers like spikes or HFOs (Zijlmans *et al*., 2012; Frauscher *et al*., 2017; Fitzgerald *et al*., 2021; Tamilia *et al*., 2021; Thomas *et al*., 2023), the considered approach offers a network-based quantitative framework that is unbiased by visual interpretation and highly robust to noise artifacts in the recordings. While fragility-related measures were typically high in clinically defined SOZ regions (Li *et al*., 2021), they were sometimes high also outside the SOZ, highlighting additional potentially unstable nodes. These might represent previously unrecognized regions of network involvement or even targets that could be used for neuromodulation in multifocal epilepsies.

In the subset of patients who underwent 50 Hz stimulation mapping, we found significant correlations between the stimulation-evoked changes in epileptiform discharges and the fragility-based metrics, reinforcing the interpretation of fragility as a proxy for network responsiveness to external perturbation. While the clinical scoring of stimulation responses (Trébuchon and Chauvel, 2016; Ritaccio, Brunner and Schalk, 2018; Grande, Ihnen and Arya, 2020) is often limited by subjectivity and poor granularity, model-based network fragility offers a quantitative and scalable complement that could assist in prioritizing electrode contacts for stimulation. In large-scale implantations with more than 10 electrodes and up to 100 contacts, exhaustive stimulation becomes time-wise unfeasible. Thus, model nodes that have been identified as drivers of network instability could guide the selection of priority contacts for targeted stimulation.

We hypothesized that the removal of instability-driving network nodes would lead to significantly higher resilience to disruption (lower fragility) in the remaining network, while removal of noninvolved or misidentified nodes would have negligible impact. To test this, we used model-based virtual thermocoagulation to evaluate how network fragility changes after virtually removing nodes that had been coagulated based on clinical SOZ identification. Our finding that in some patients the resulting network became significantly more resilient suggests that brain tissue around the thermocoagulated channels had a genuine destabilizing role. In other patients, the network-level effect was weak or absent, highlighting inter-patient variability and potential limitations of local interventions. Interestingly, in almost all cases, the most prominent changes were located near the removed nodes, which might reflect the underlying neuroanatomy and, consequently, stronger connections in the network model.

Several limitations should be acknowledged. The definition of the SOZ against which all metrics are compared is based on clinical agreement and may not always be precise (Jayakar *et al*., 2016; Chowdhury *et al*., 2021). Responses to 50 Hz stimulation are difficult to evaluate in patients with very active interictal backgrounds, where frequent epileptiform discharges occur even without stimulation. Additionally, even though high fragility may suggest susceptibility to pathological state transitions, in principle, network instability could also lead to functional or healthy dynamic states, as seen in eloquent cortex (John and Ahmed, 2022). Therefore, weak or nonsignificant correlations with the clinical SOZ or stimulation response are not necessarily unexpected and do not reduce the potential utility of the metrics.

A significant difference in metric values between SOZ and non-SOZ channels does not automatically imply that channels can be reliably classified in a prospective way. Such interpretations are often confounded when combined with machine learning pipelines, where model performance depends on choice of classifying algorithms and where summary metrics like the area under the receiver operating curve (AUC-ROC) may mask underlying uncertainty. To avoid this limitation, we intentionally did not include classification models or AUC analyses, as they can overstate per-channel predictive utility. Despite this, higher fragility or source-sink values indicate increased probability of SOZ involvement and can inform clinical decisions, such as selecting channels for stimulation during presurgical evaluation.

The considered brain areas are not isolated systems and still receive inputs from unrecorded regions, which are not captured in our models. Electrode contacts may also lie in structurally or functionally distinct tissue types, including white matter or subcortical regions (Bernabei *et al*., 2023). Nevertheless, we showed that by including these areas and using a limited set of channels clinically meaningful results can still be achieved. A more detailed analysis of how fragility and related measures depend on tissue type and anatomical location would be an important direction for future work.

Longer-term outcomes after thermocoagulation could not be systematically assessed, as most patients subsequently underwent resective surgery. A larger cohort of patients undergoing thermocoagulation alone, or a cohort whose resected brain areas are precisely mapped to the initial locations of sEEG contacts, would be valuable to evaluate whether predicted increases in network resilience correlate with clinical outcome.

Technically, the extremely high similarity of incoming and outgoing fragility, as well as of source and sink metrics, is largely due to the estimated network matrices being close to symmetric. This also explains why the at-first unintuitive definition of sink connectivity, as the influence from other sink-like nodes rather than to them, still yields meaningful results, since *A*_*ij*_ ≈ *A*_*ji*_. Whether this holds at longer timescales or larger spatial scales remains an open question, but it consistently arises from fitting short-term local dynamics. The absence of frequency dependence in fragility indicates that the network dynamics we estimate do not have any mode that is especially close to instability at a particular oscillatory frequency. Instead, all eigenmodes of *A* appear similarly stable, which is consistent with the effect of ridge regularization in our model estimation (see Methods), where all modes are pulled away from the instability boundary by a similar amount. In this situation, changing the frequency parameter in the fragility computation has little effect, because all modes are equally hard to destabilize. Differences in fragility between channels therefore reflect only how strongly each channel participates in these eigenmodes, as determined by the network’s spatial structure. This also explains the high correlation of fragility with frequency-agnostic measures like the source–sink metrics, which capture similar spatial aspects of the underlying dynamics. The stability of results across frequency, time, and metric variants suggests that the fragile nodes we identify are determined mainly by the network’s spatial organization, rather than by resonance with specific brain rhythms.

In summary, our results support the feasibility and clinical relevance of model-based interictal network analysis. The methods are compatible with existing sEEG recordings and offer interpretable, patient-specific insights that can guide SOZ identification, improve the understanding of stimulation effects during presurgical evaluation, and support virtual resection planning.

## Supporting information

Dynamic network-based metrics for all included patients.

Comparison of network-based metrics and reactions to 50 Hz stimulation for all included patients.

## Data Availability

All data produced in the present study are available upon reasonable request to the authors.

## Acknowledgement

We are grateful to Matija Varga and Marc Serra Garcia for inspiring discussions. This work was funded by the Swiss National Science Foundation grant (SNF 197766, awarded to L.I and R.P.), by the European Research Council (ERC) under the European Union’s Horizon 2020 research and innovation program (grant agreement no. 758604), by an ERC starting grant (ENTRAINER, awarded to R.P.), by an ETH Grant (ETH-25 18-2, awarded to R.P.), by the Koetser Foundation research grant (awarded to T.D and L.I.) and by the research grants from the Swiss Epilepsy Foundation.

