## Supplementary material for "Bridging Computational and Clinical Strategies to Improve Presurgical Identification of Epileptogenic Networks": Dynamic network-based metrics for all included patients.

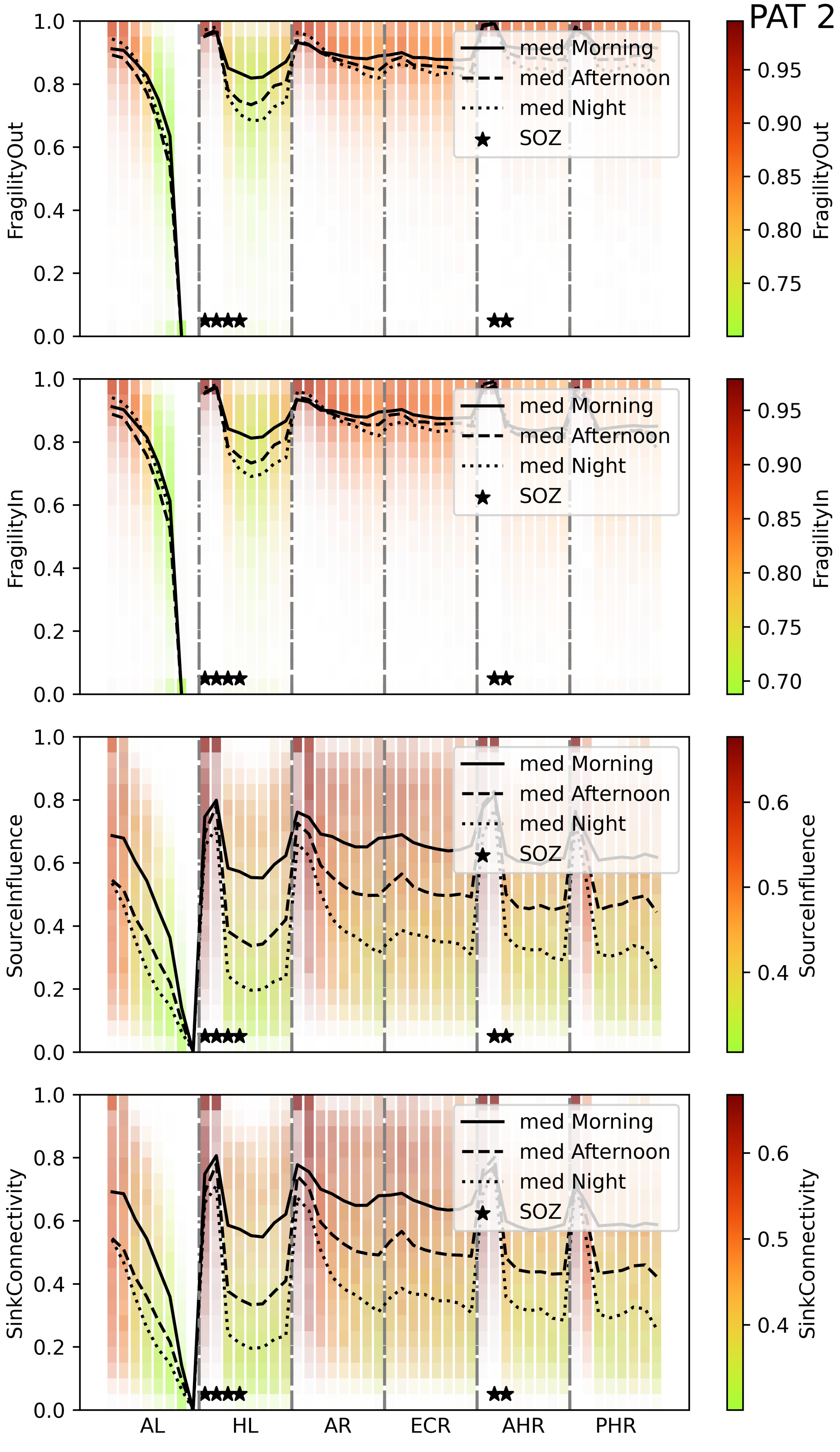
PAT 2
FragilityOut
FragilityIn
SourceInfluence
SinkConnectivity
med Morning
med Afternoon
med Night
SOZ
AL
HL
AR
ECR
AHR
PHR

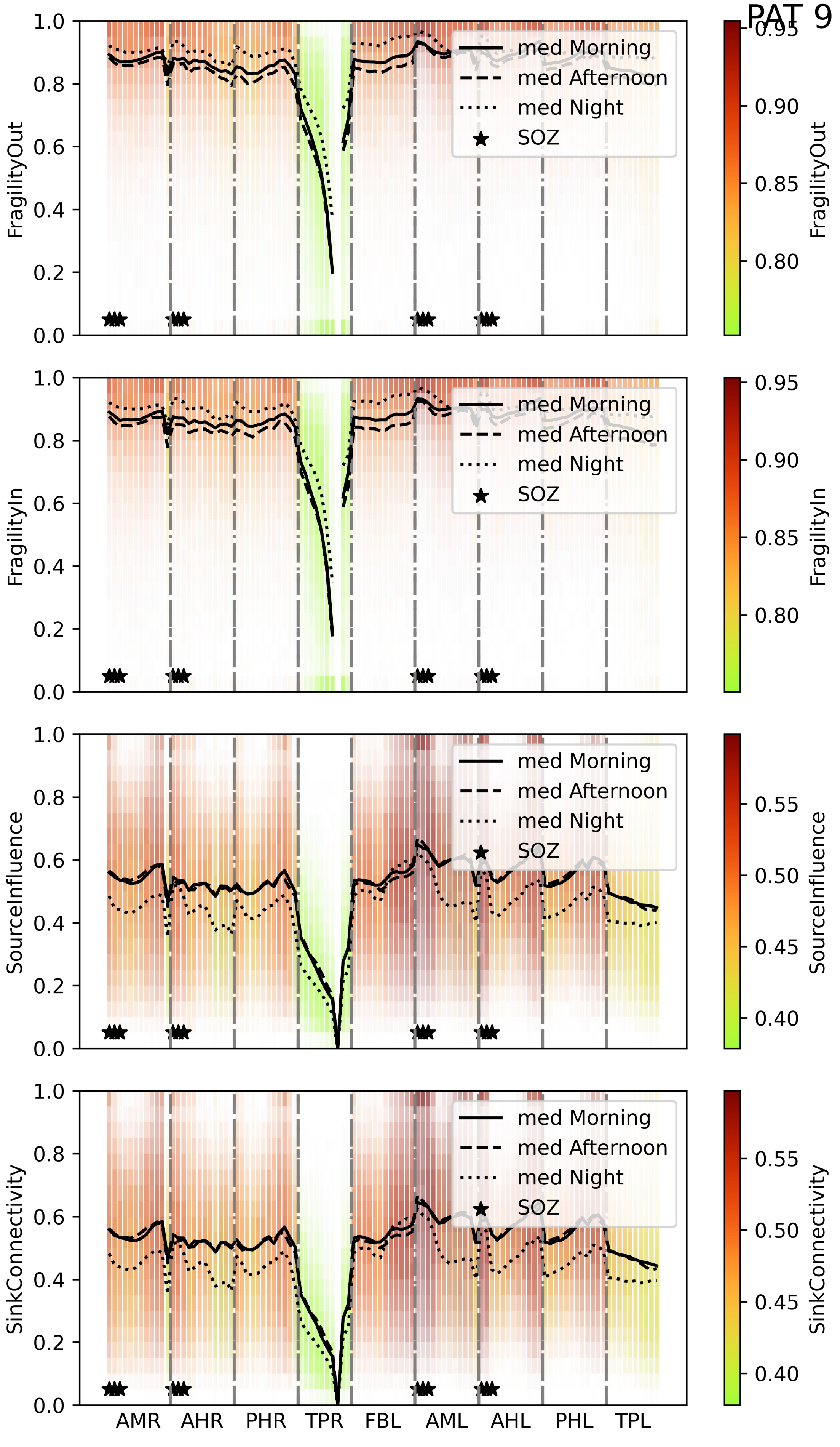
PAT 9
FragilityOut
FragilityIn
SourceInfluence
SinkConnectivity
med Morning
med Afternoon
med Night
SOZ
1.0
0.8
0.6
0.4
0.2
0.0
0.95
0.90
0.85
0.80
0.55
0.50
0.45
0.40
AMR
AHR
PHR
TPR
FBL
AML
AHL
PHL
TPL

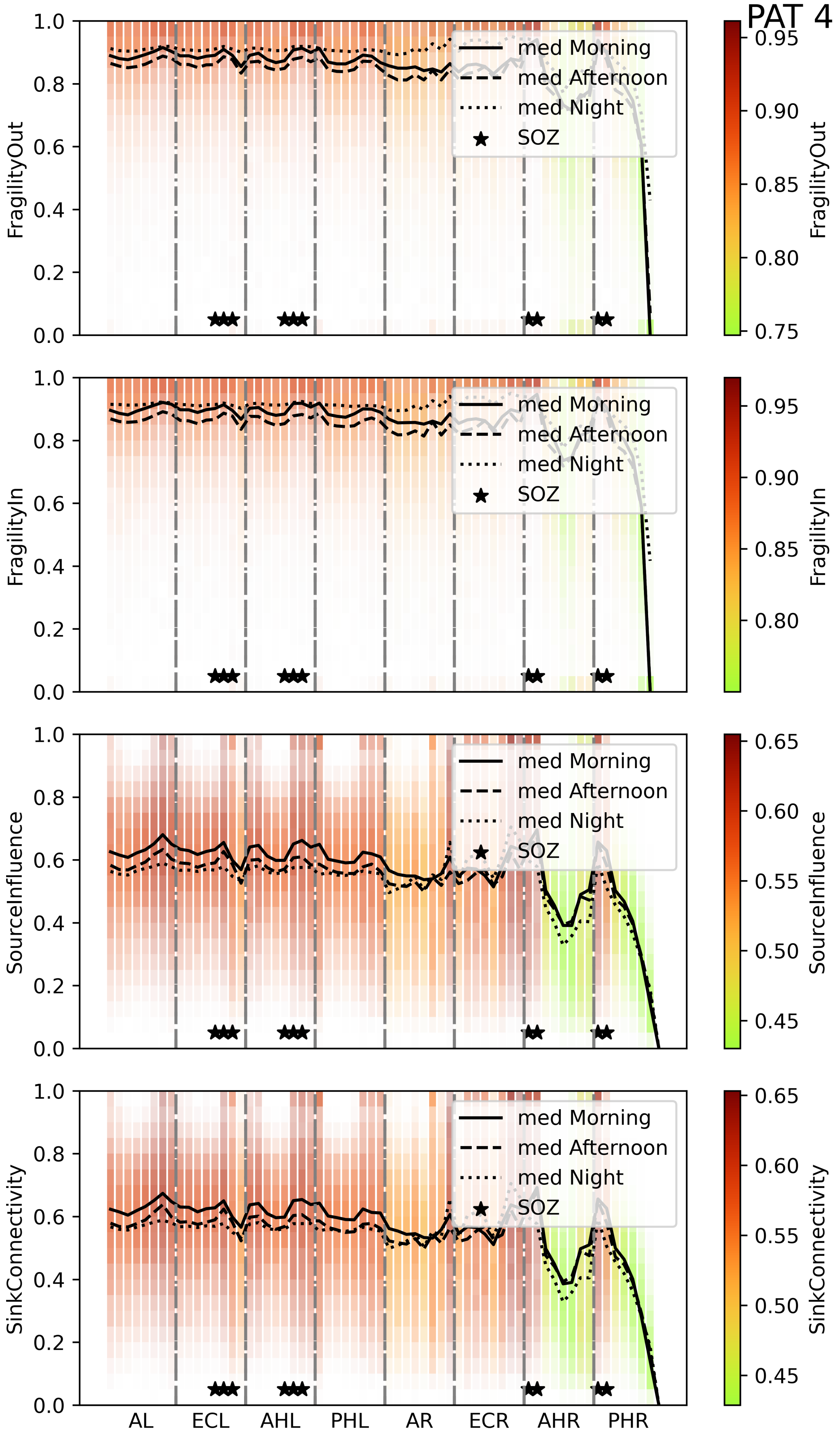
PAT 4
FragilityOut
FragilityIn
SourceInfluence
SinkConnectivity
med Morning
med Afternoon
med Night
SOZ
AL
ECL
AHL
PHL
AR
ECR
AHR
PHR

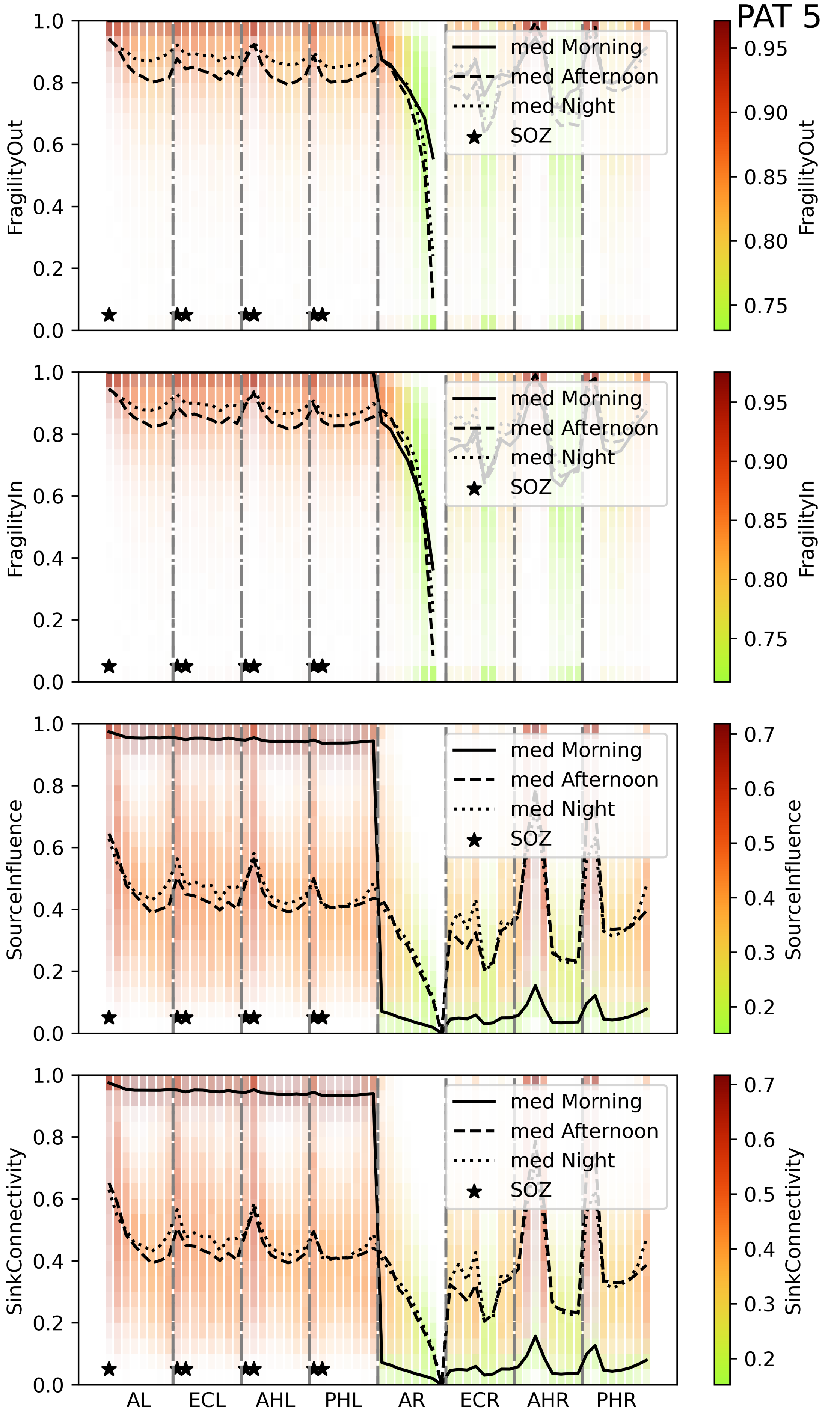
PAT 5
FragilityOut
FragilityIn
SourceInfluence
SinkConnectivity
med Morning
med Afternoon
med Night
SOZ
AL
ECL
AHL
PHL
AR
ECR
AHR
PHR

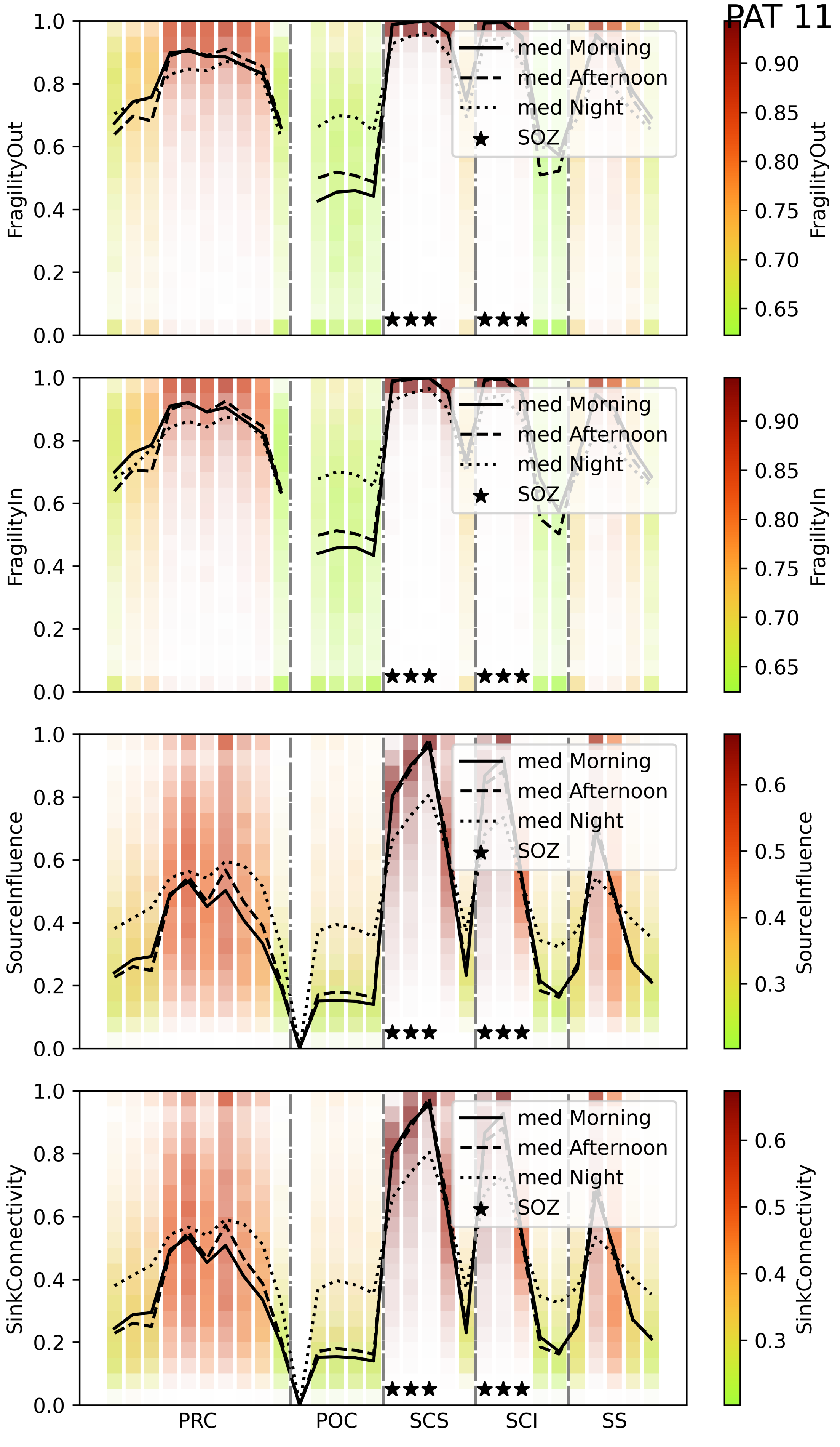
PAT 11
FragilityOut
FragilityIn
SourceInfluence
SinkConnectivity
med Morning
med Afternoon
med Night
SOZ
PRC
POC
SCS
SCI
SS

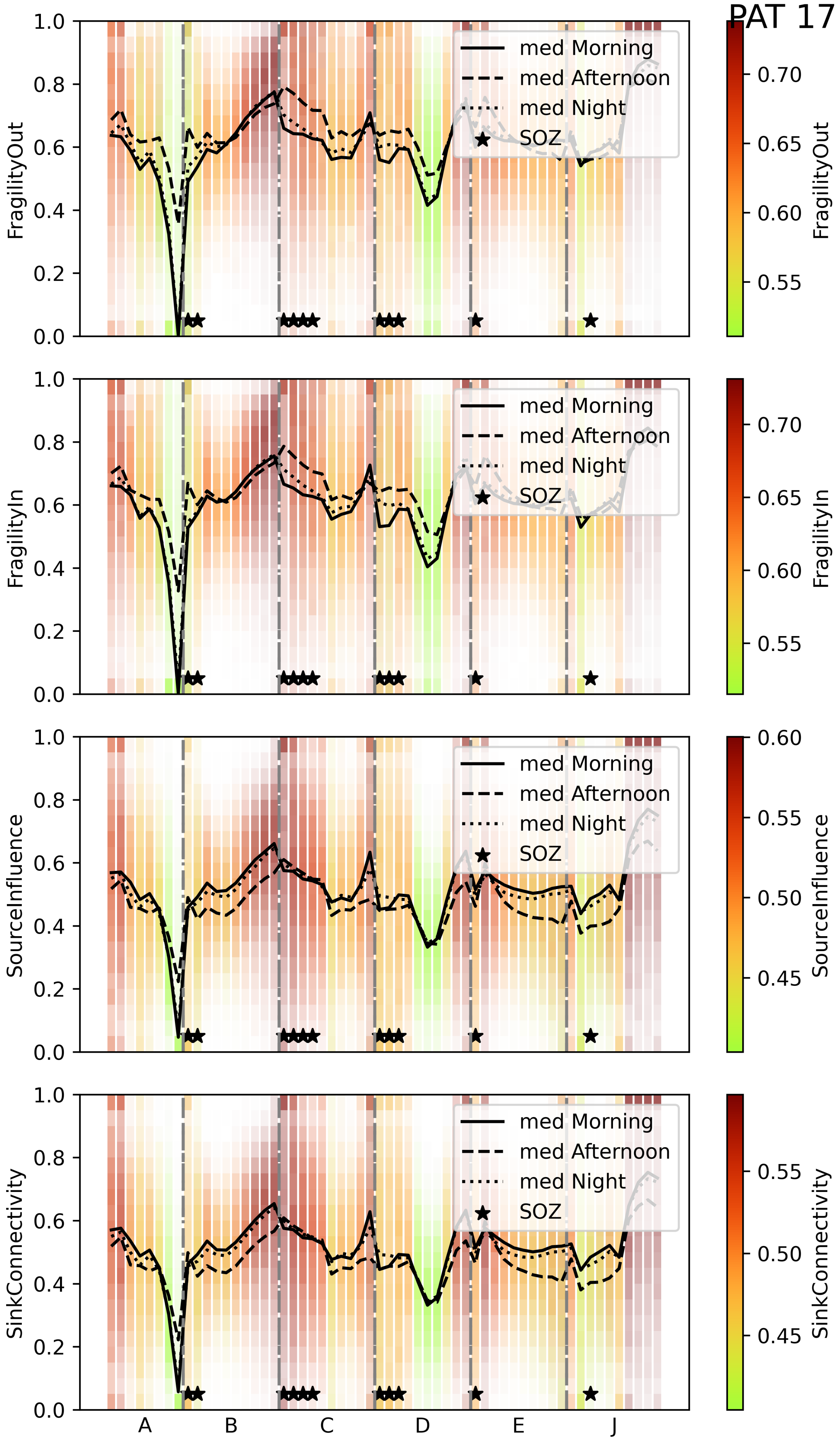
PAT 17
FragilityOut
FragilityIn
SourceInfluence
SinkConnectivity
med Morning
med Afternoon
med Night
SOZ
A
B
C
D
E
J

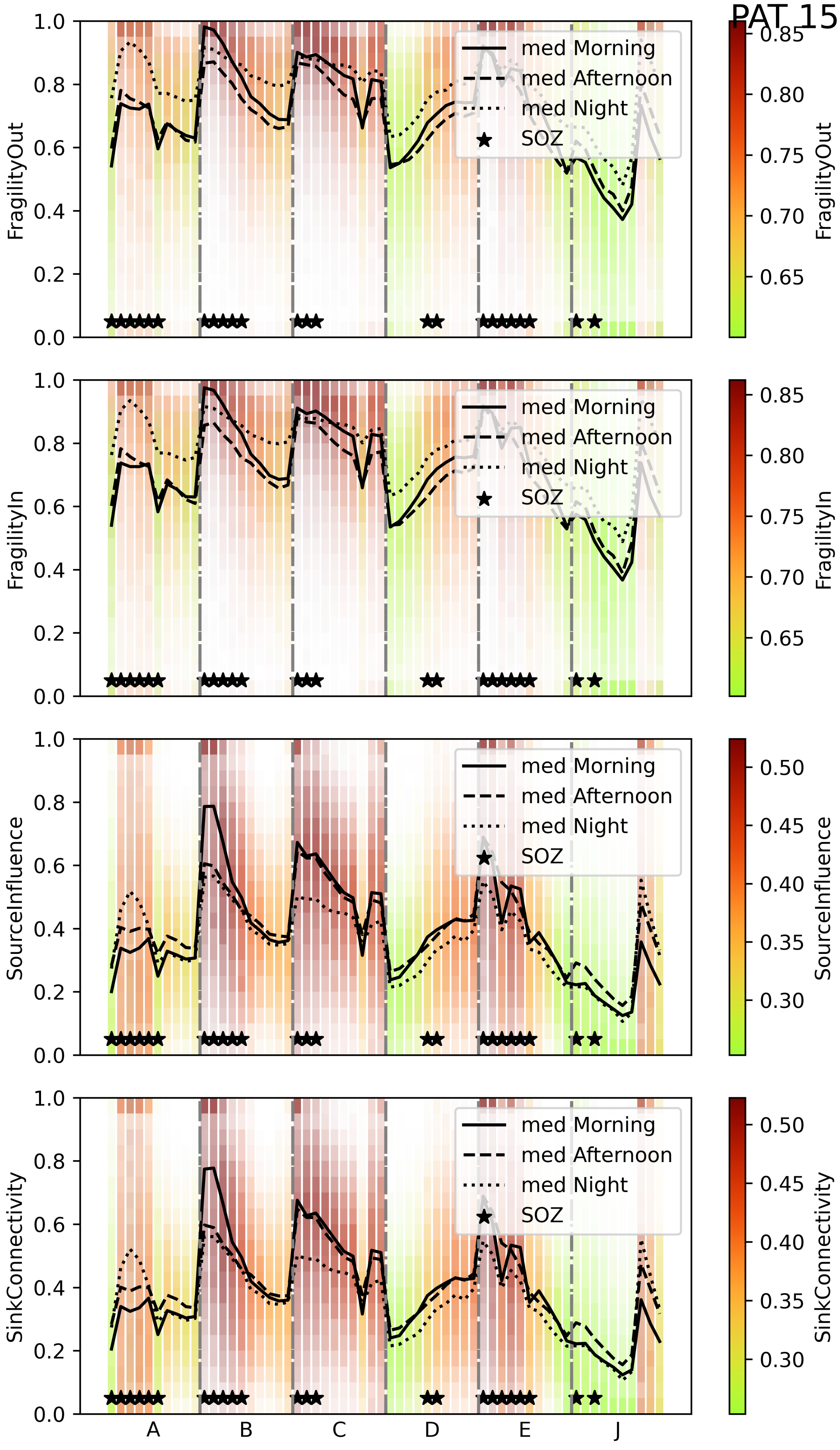

PAT 15
FragilityOut
FragilityIn
SourceInfluence
SinkConnectivity
med Morning
med Afternoon
med Night
SOZ
1.0
0.8
0.6
0.4
0.2
0.0
0.85
0.80
0.75
0.70
0.65
0.50
0.45
0.40
0.35
0.30
A
B
C
D
E
J

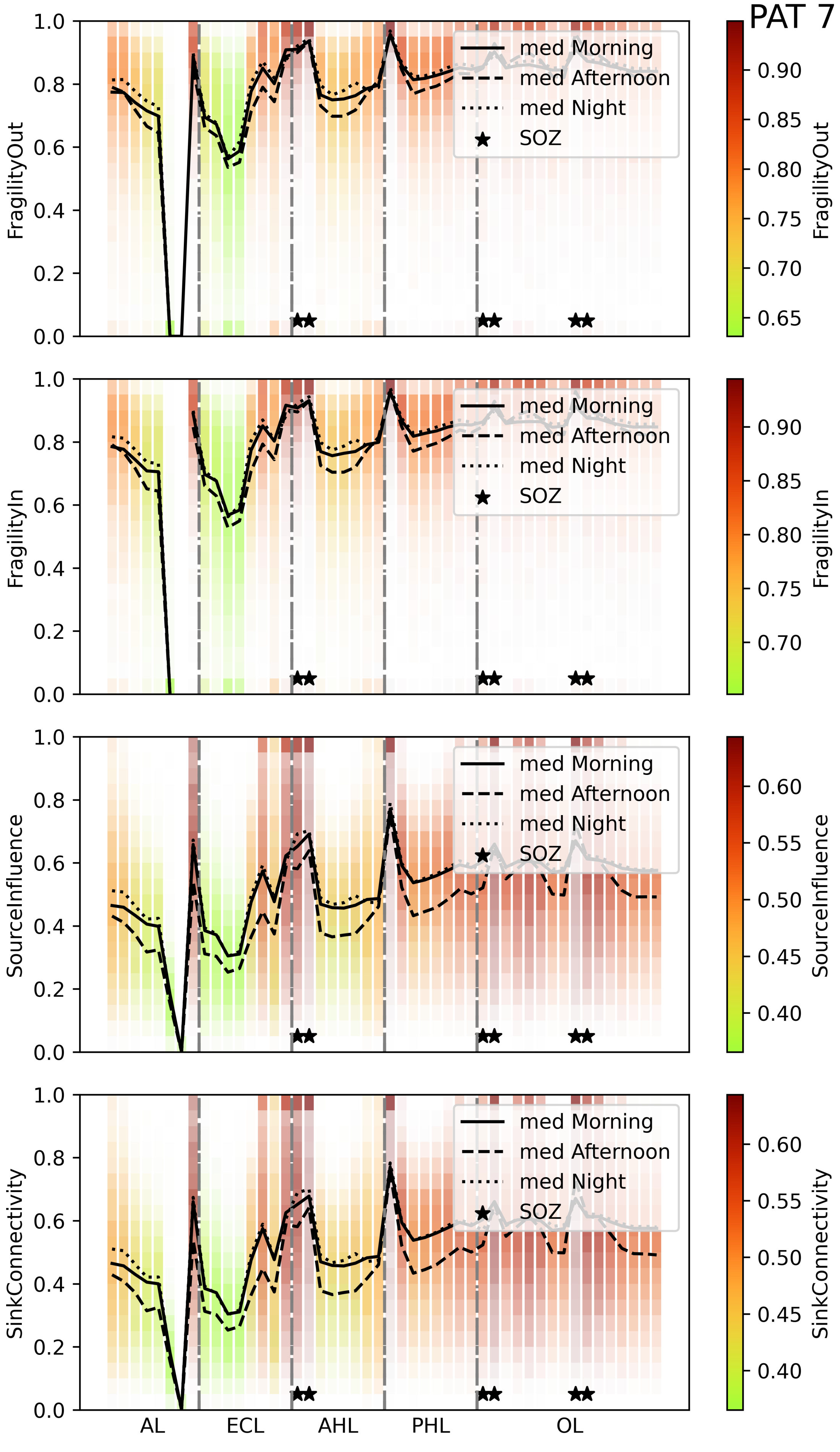

PAT 7
FragilityOut
med Morning
med Afternoon
med Night
SOZ
FragilityIn
SourceInfluence
SinkConnectivity
AL
ECL
AHL
PHL
OL

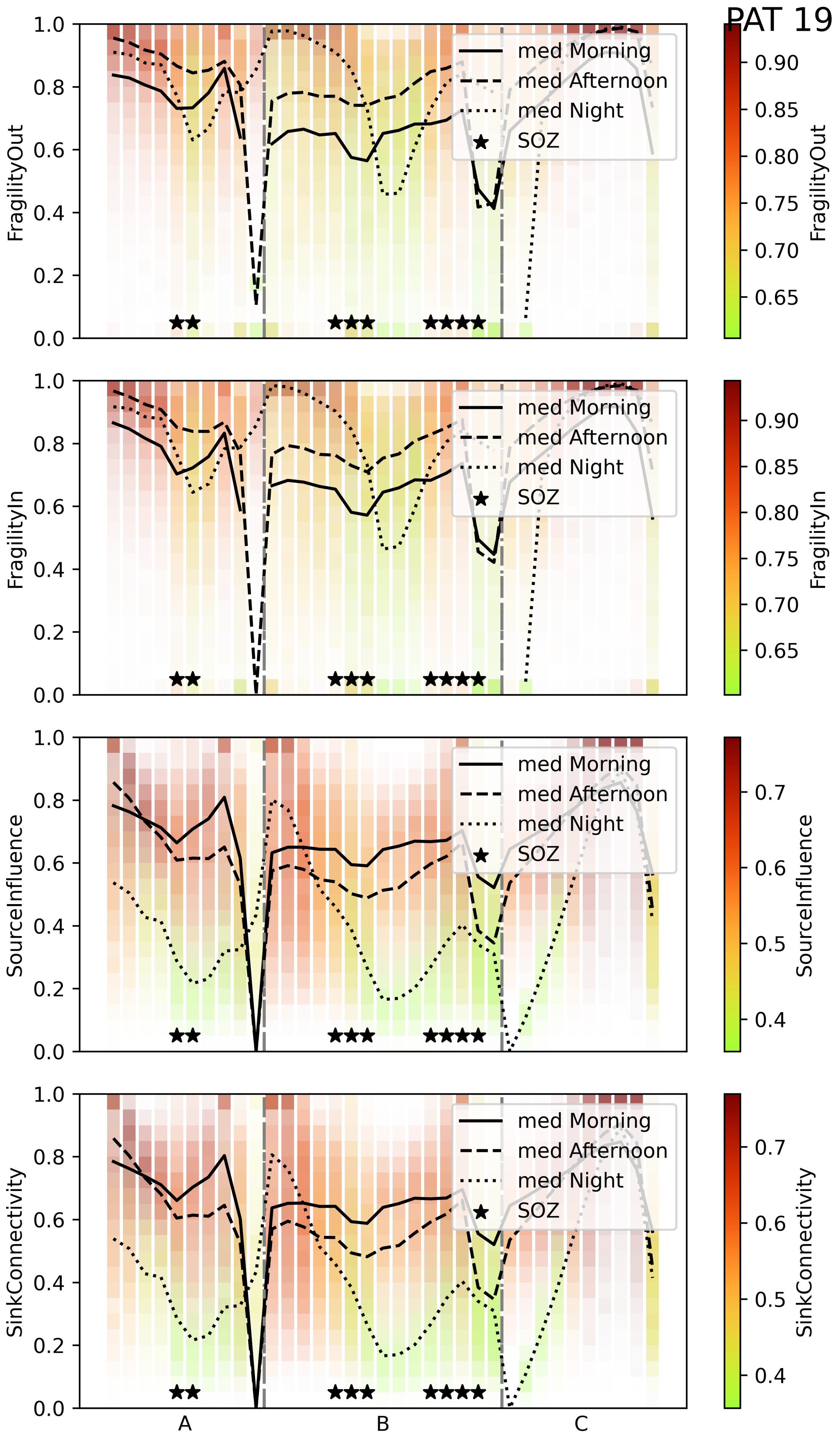
PAT 19
FragilityOut
FragilityIn
SourceInfluence
SinkConnectivity
med Morning
med Afternoon
med Night
SOZ
A
B
C
0.0
0.2
0.4
0.6
0.8
1.0
0.65
0.70
0.75
0.80
0.85
0.90
0.4
0.5
0.6
0.7

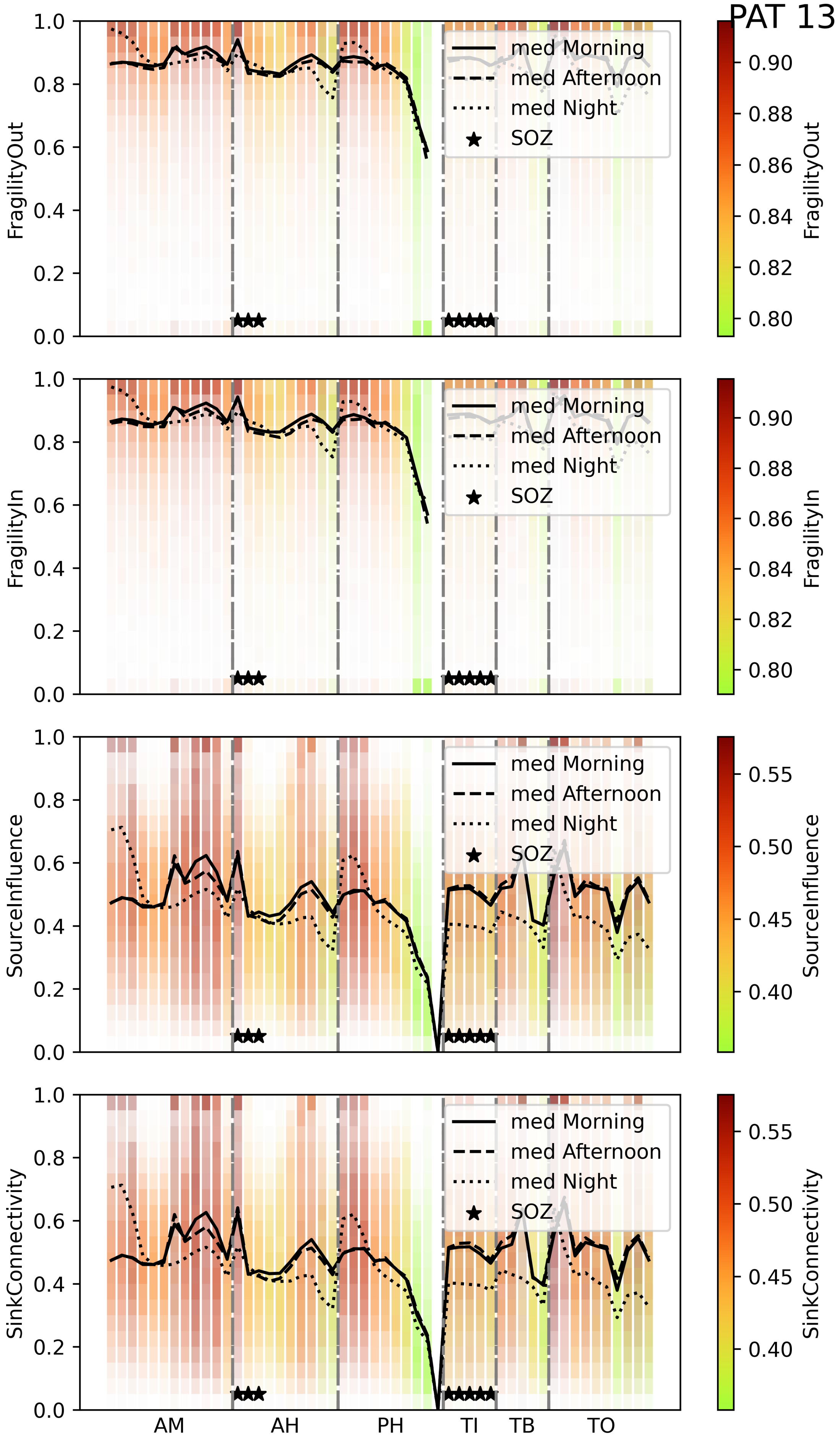
PAT 13
FragilityOut
FragilityIn
SourceInfluence
SinkConnectivity
med Morning
med Afternoon
med Night
SOZ
AM
AH
PH
TI
TB
TO

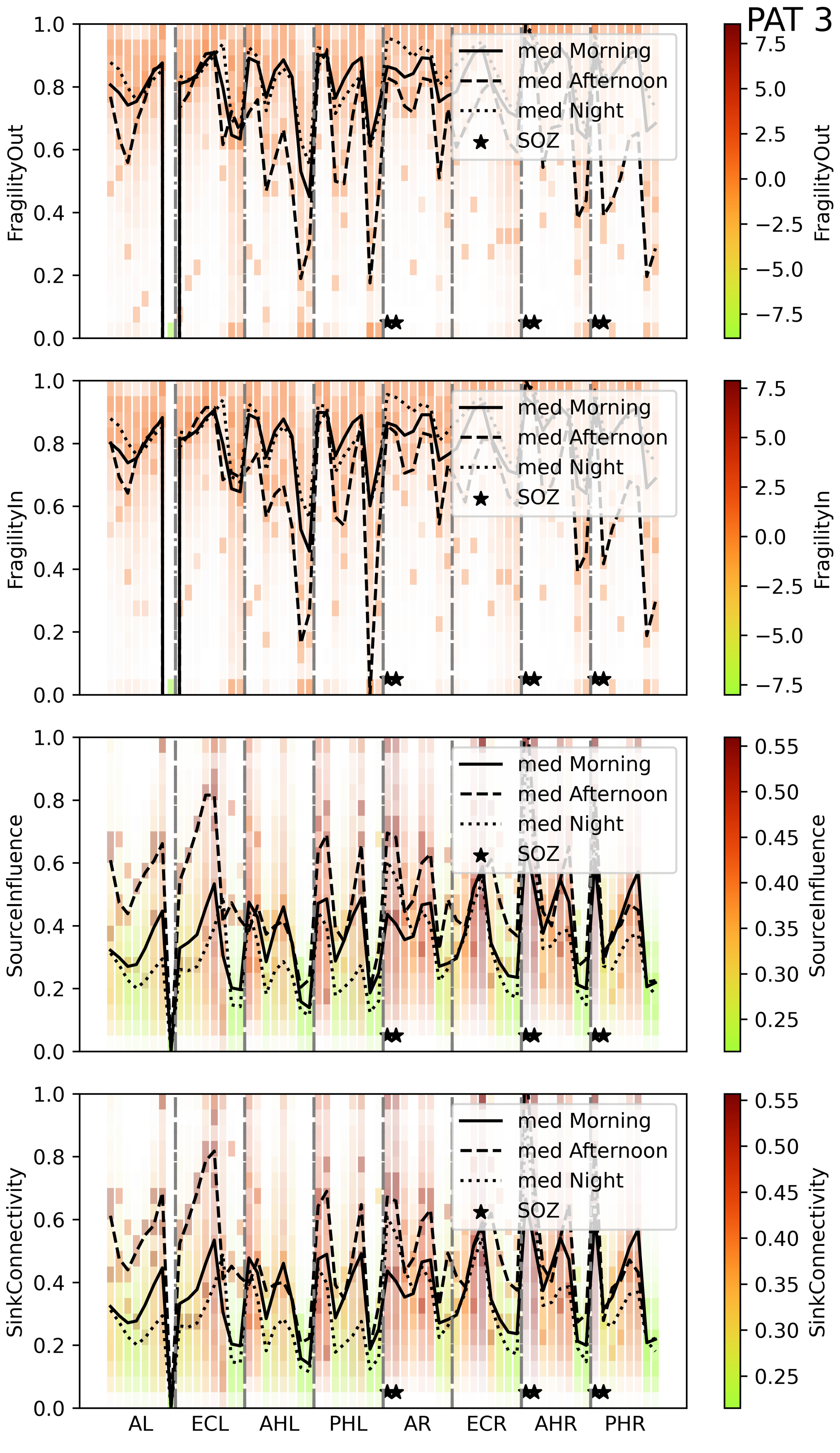

PAT 3
FragilityOut
FragilityIn
SourceInfluence
SinkConnectivity
med Morning
med Afternoon
med Night
SOZ
AL
ECL
AHL
PHL
AR
ECR
AHR
PHR

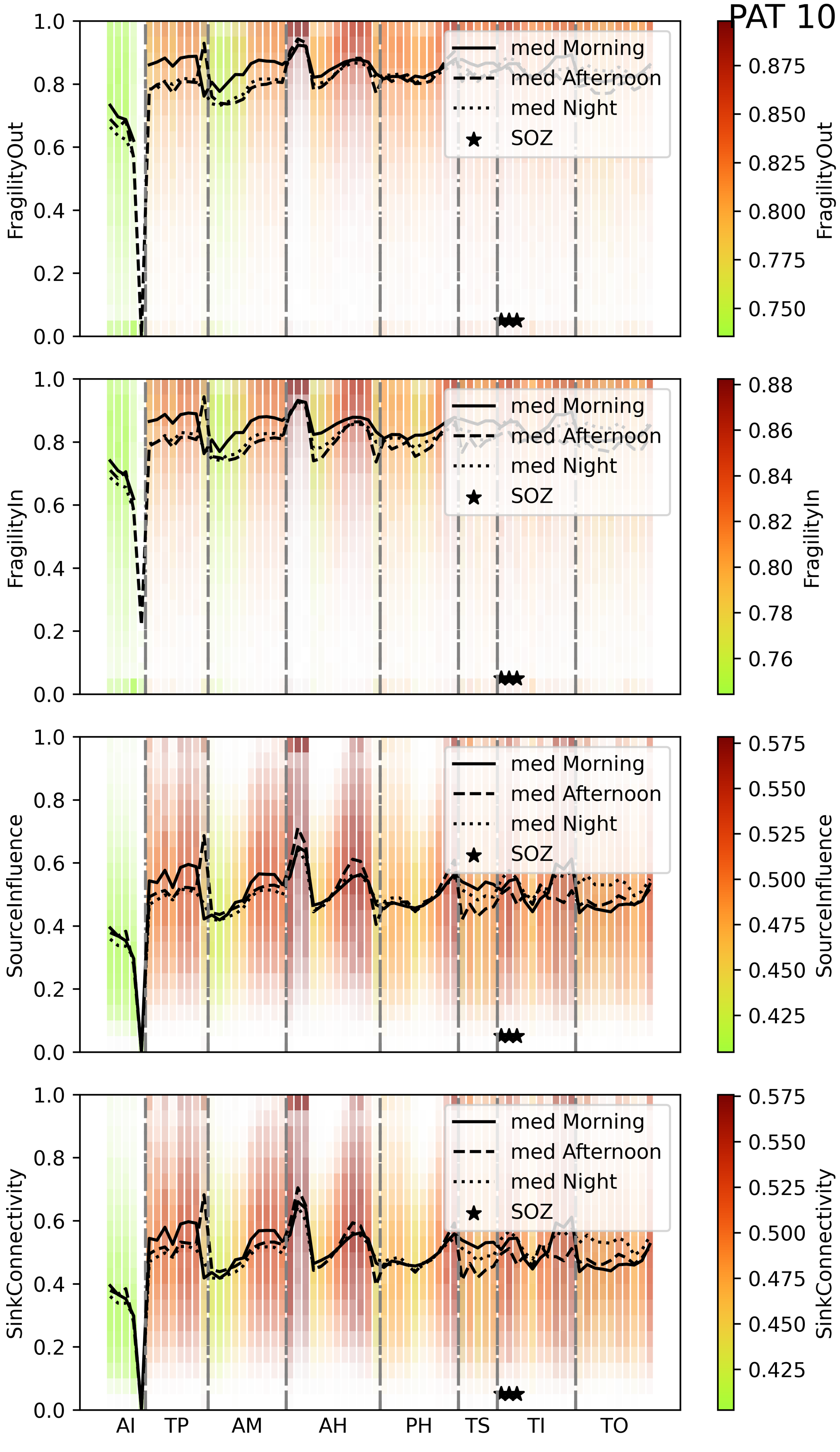
PAT 10
FragilityOut
FragilityIn
SourceInfluence
SinkConnectivity
med Morning
med Afternoon
med Night
SOZ
AI
TP
AM
AH
PH
TS
TI
TO

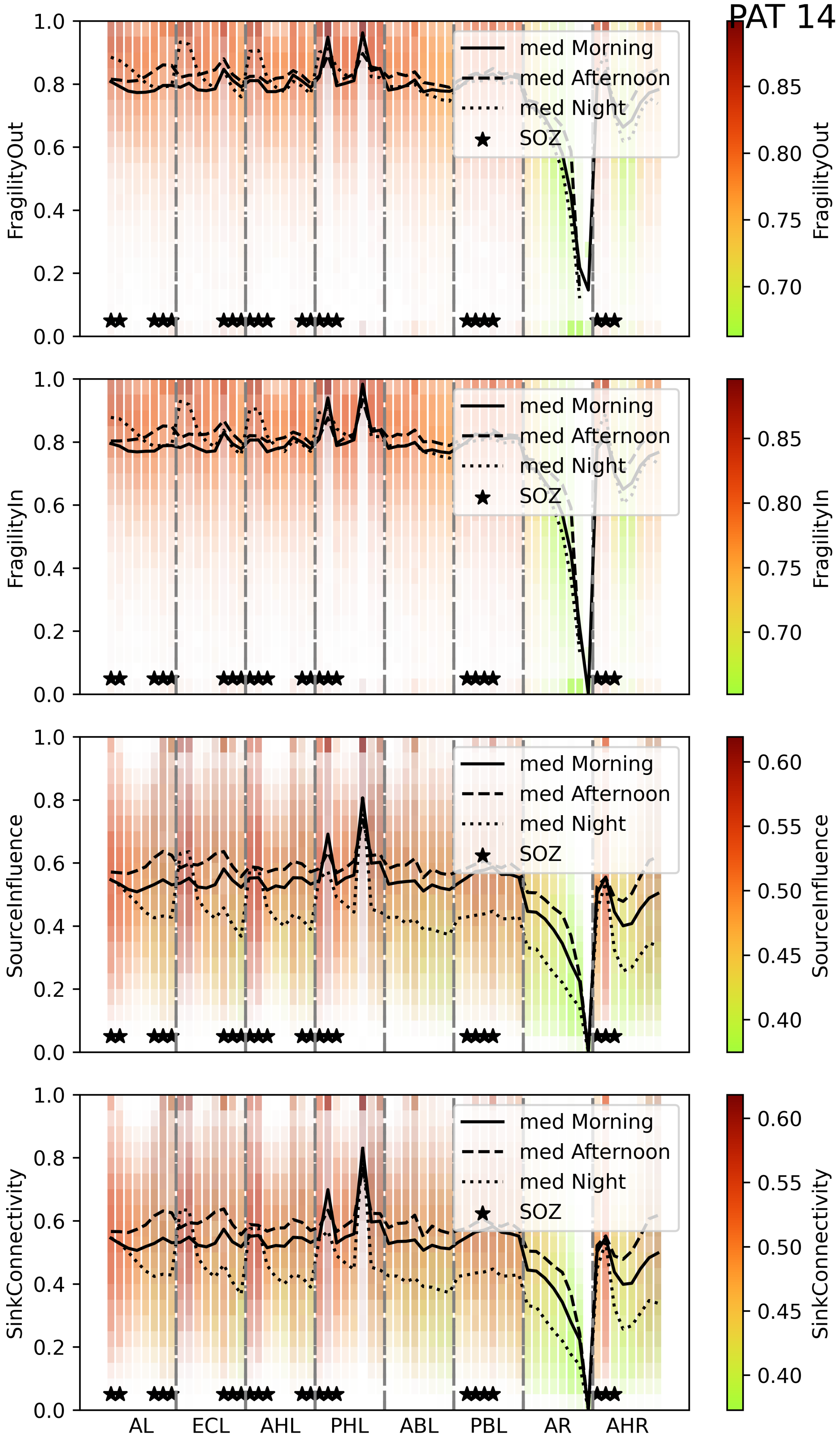
PAT 14
FragilityOut
FragilityIn
SourceInfluence
SinkConnectivity
med Morning
med Afternoon
med Night
SOZ
AL
ECL
AHL
PHL
ABL
PBL
AR
AHR

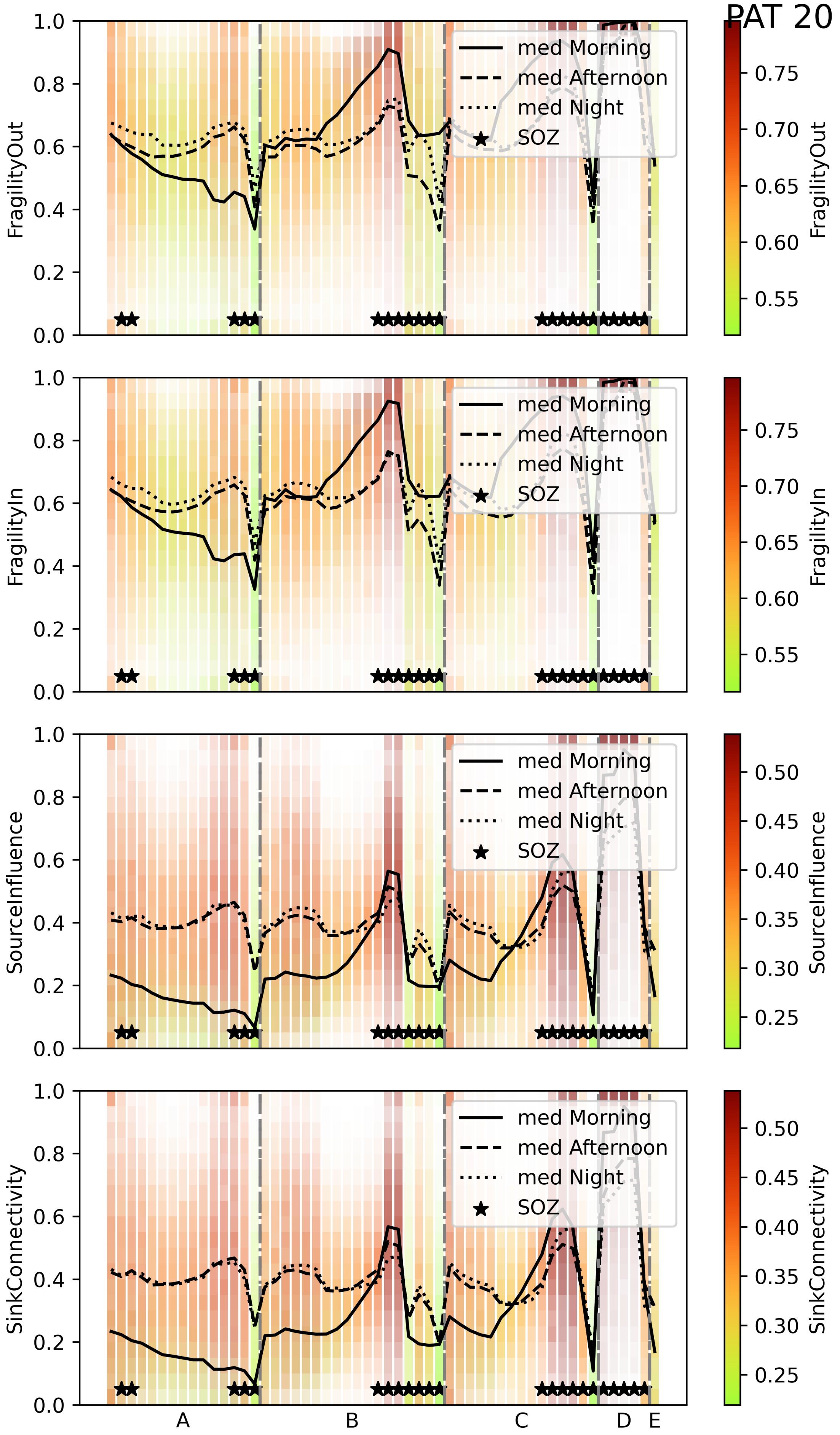
PAT 20
FragilityOut
FragilityIn
SourceInfluence
SinkConnectivity
med Morning
med Afternoon
med Night
SOZ
A
B
C
D
E

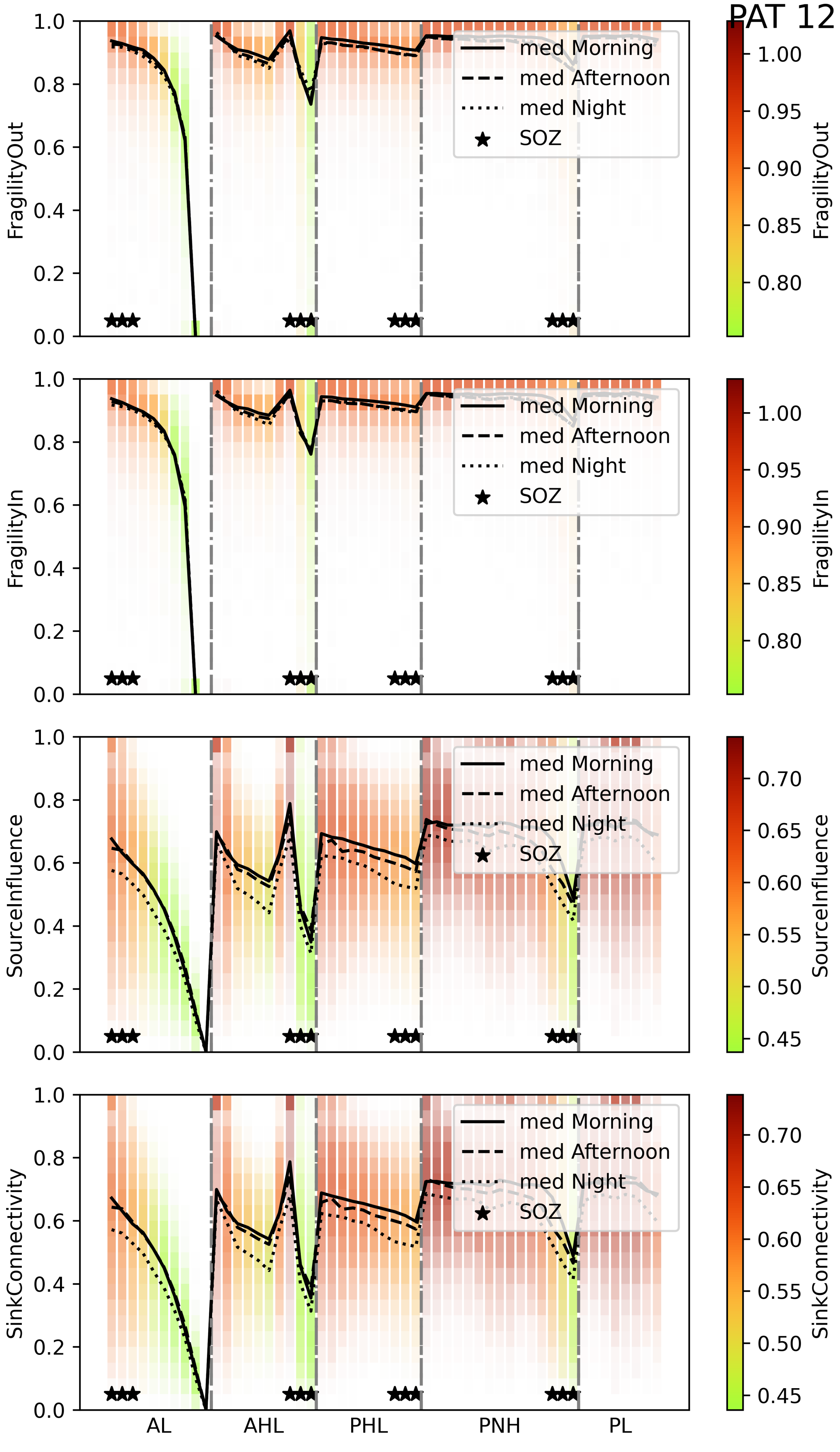
PAT 12
FragilityOut
FragilityIn
SourceInfluence
SinkConnectivity
med Morning
med Afternoon
med Night
SOZ
AL
AHL
PHL
PNH
PL
0.0
0.2
0.4
0.6
0.8
1.0
0.80
0.85
0.90
0.95
1.00
0.45
0.50
0.55
0.60
0.65
0.70

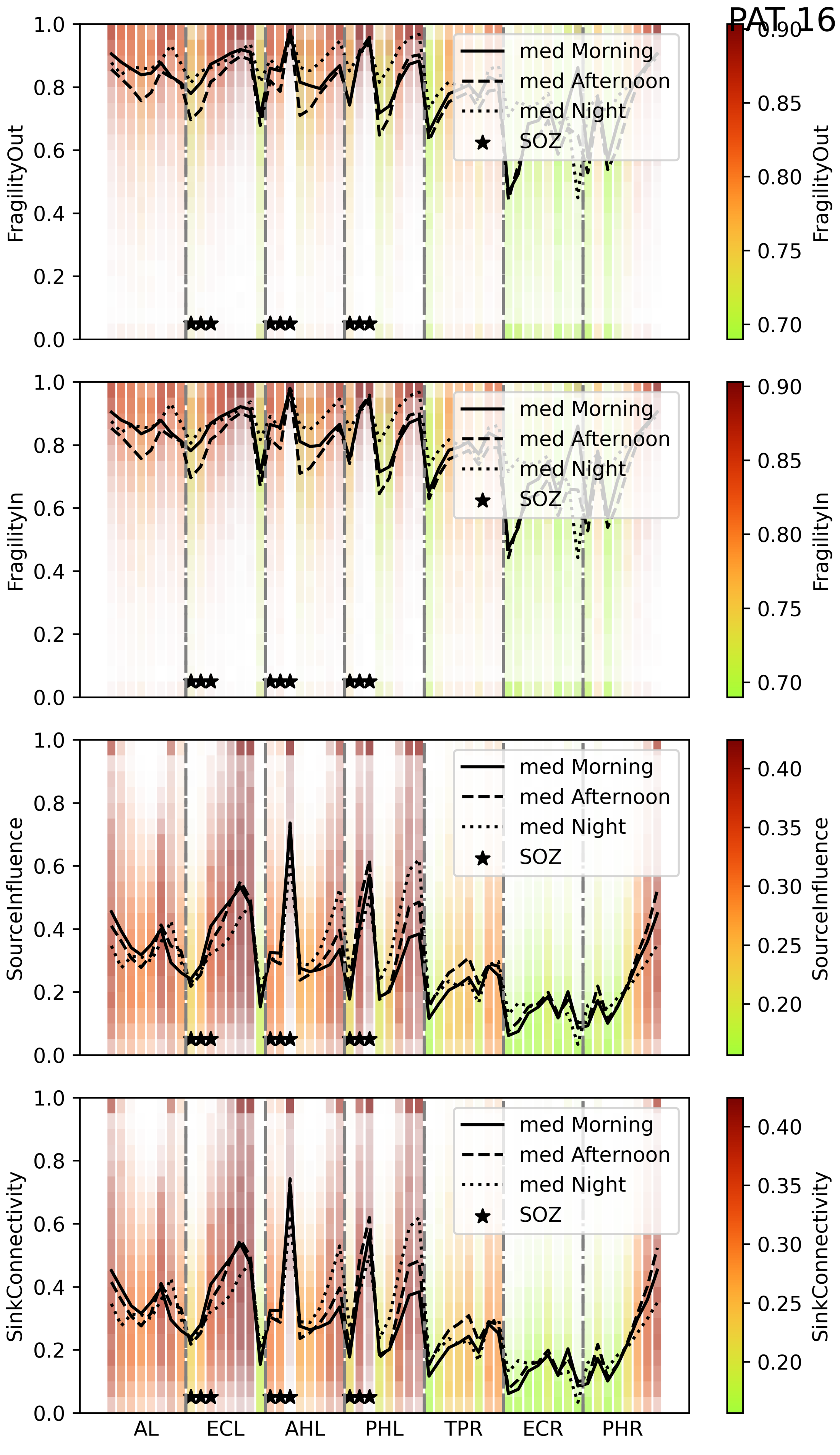

PAT 16
FragilityOut
FragilityIn
SourceInfluence
SinkConnectivity
med Morning
med Afternoon
med Night
SOZ
AL
ECL
AHL
PHL
TPR
ECR
PHR

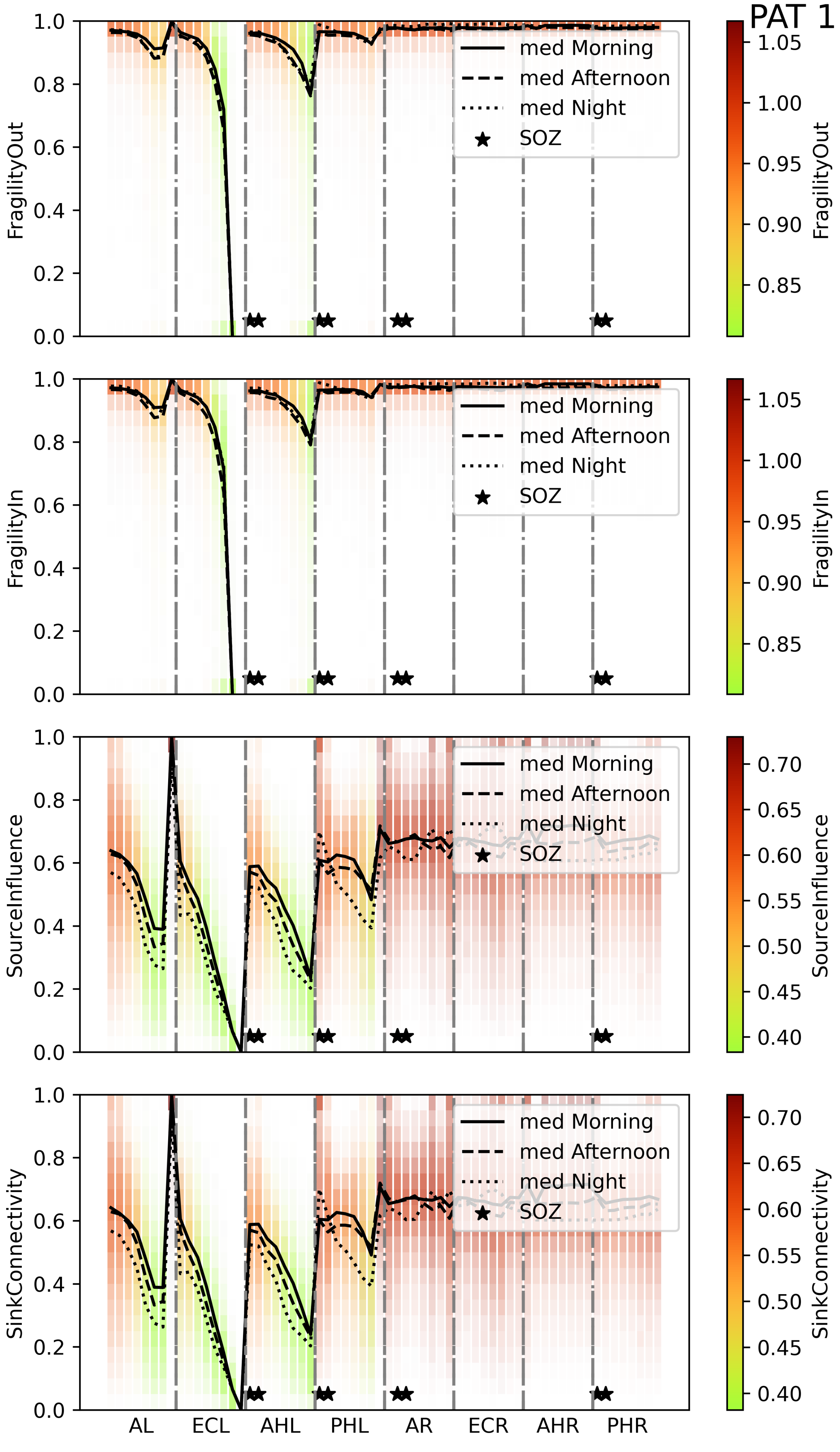
PAT 1
FragilityOut
FragilityIn
SourceInfluence
SinkConnectivity
med Morning
med Afternoon
med Night
SOZ
AL
ECL
AHL
PHL
AR
ECR
AHR
PHR

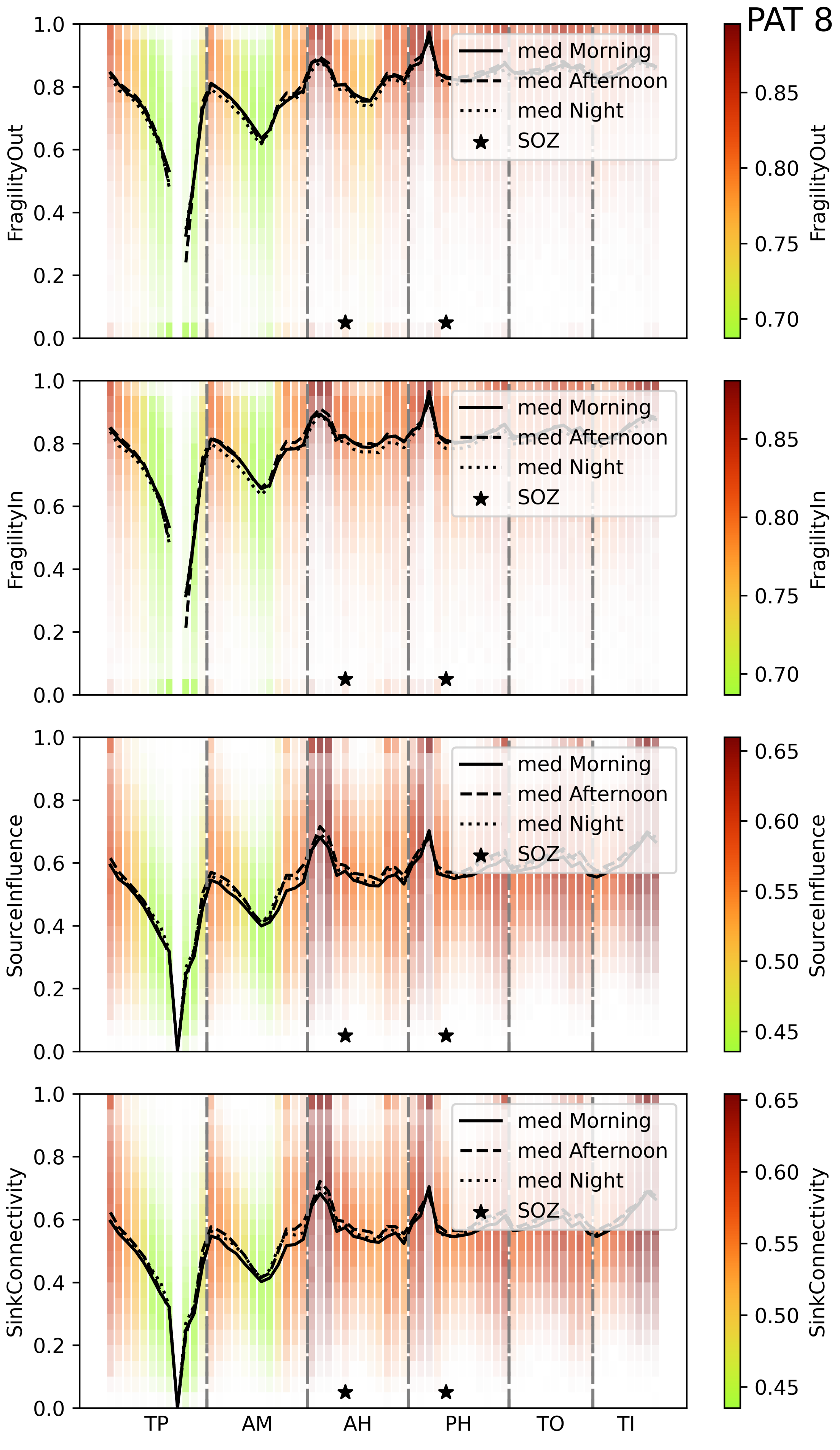
PAT 8
FragilityOut
FragilityIn
SourceInfluence
SinkConnectivity
med Morning
med Afternoon
med Night
SOZ
TP
AM
AH
PH
TO
TI
1.0
0.8
0.6
0.4
0.2
0.0
0.85
0.80
0.75
0.70
0.65
0.60
0.55
0.50
0.45

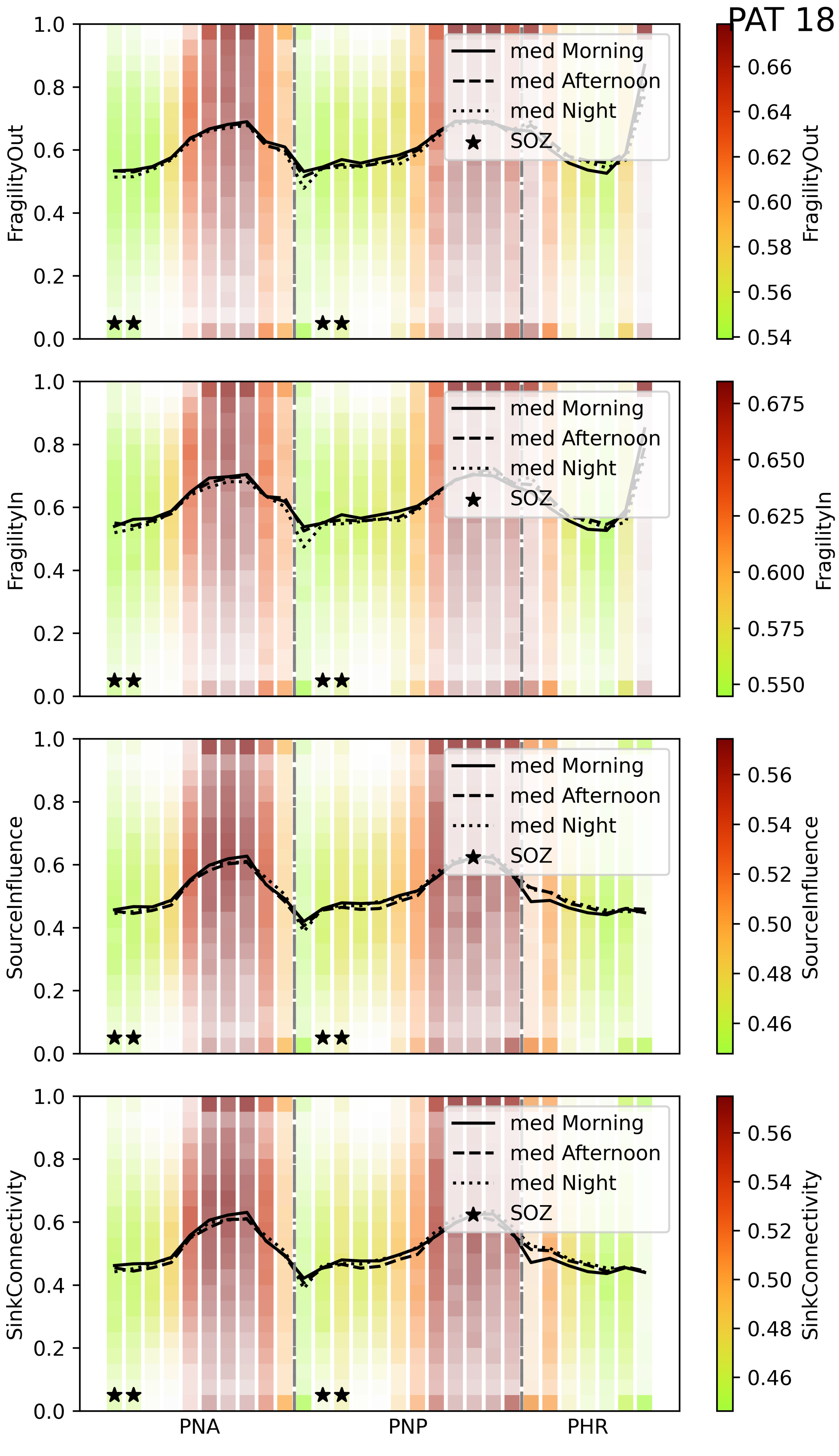
PAT 18
FragilityOut
FragilityIn
SourceInfluence
SinkConnectivity
med Morning
med Afternoon
med Night
SOZ
PNA
PNP
PHR

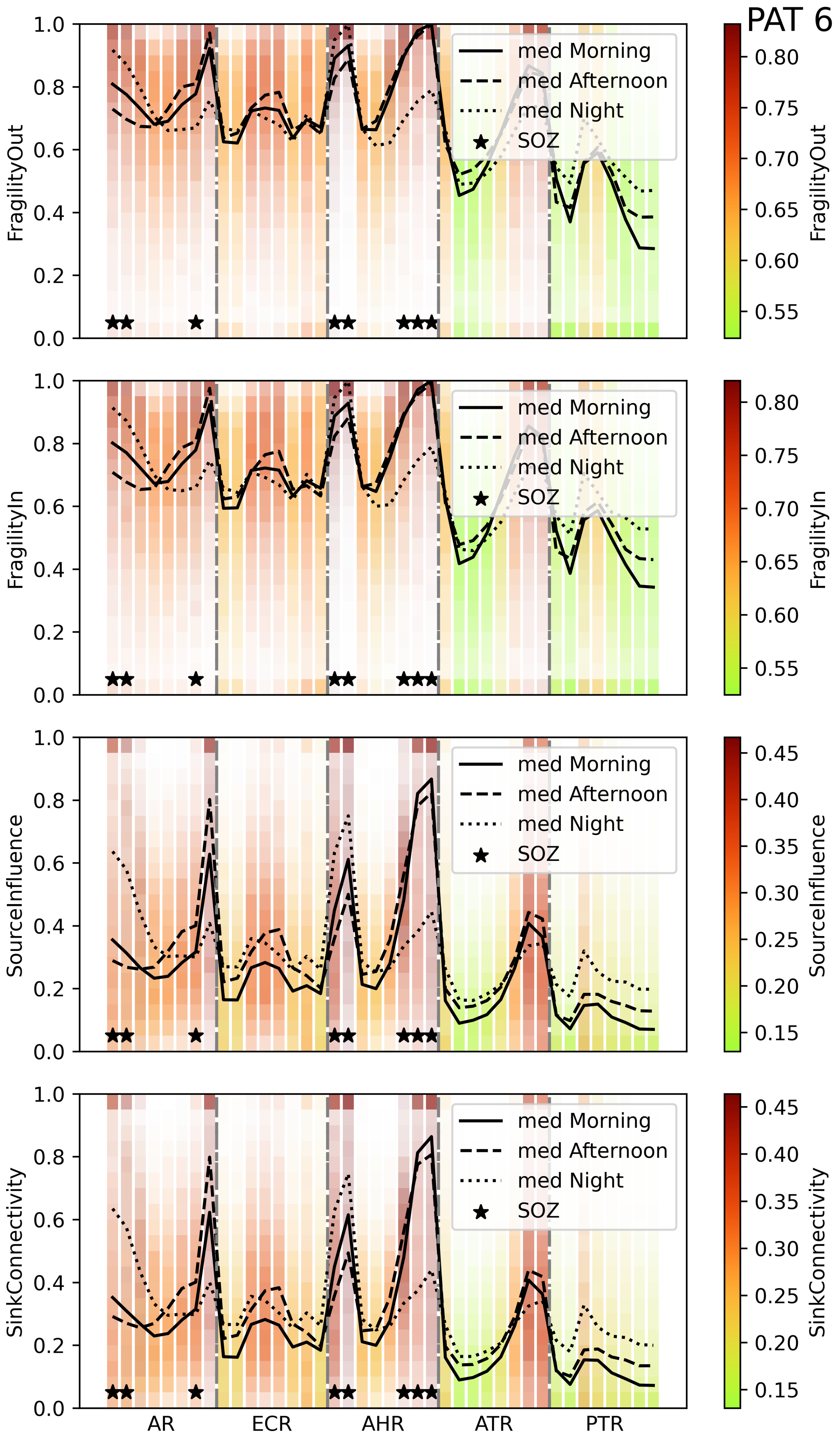
PAT 6
FragilityOut
FragilityIn
SourceInfluence
SinkConnectivity
med Morning
med Afternoon
med Night
SOZ
AR
ECR
AHR
ATR
PTR
