## Supplementary figures and images for "Bridging Computational and Clinical Strategies to Improve Presurgical Identification of Epileptogenic Networks"

### Comparison of network-based metrics and reactions to 50 Hz stimulation for all included patients.

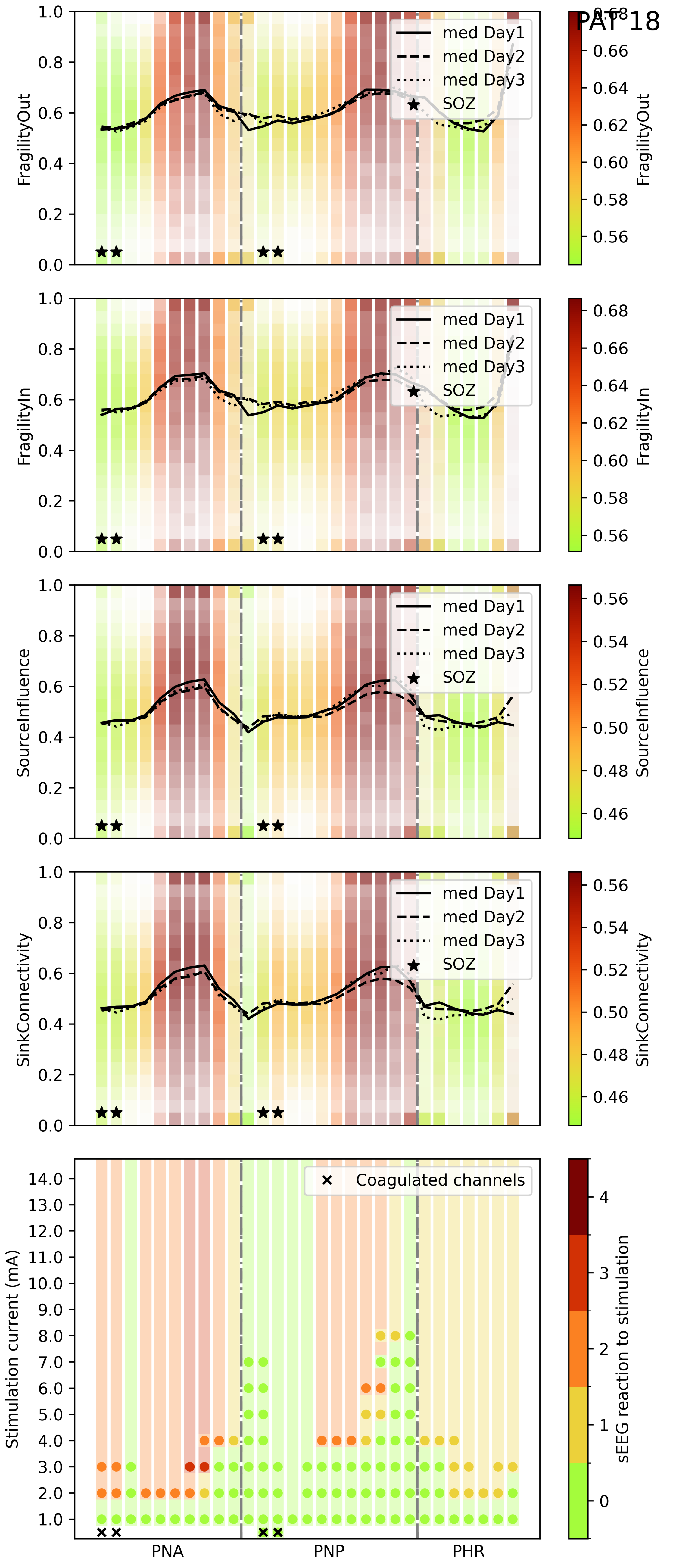

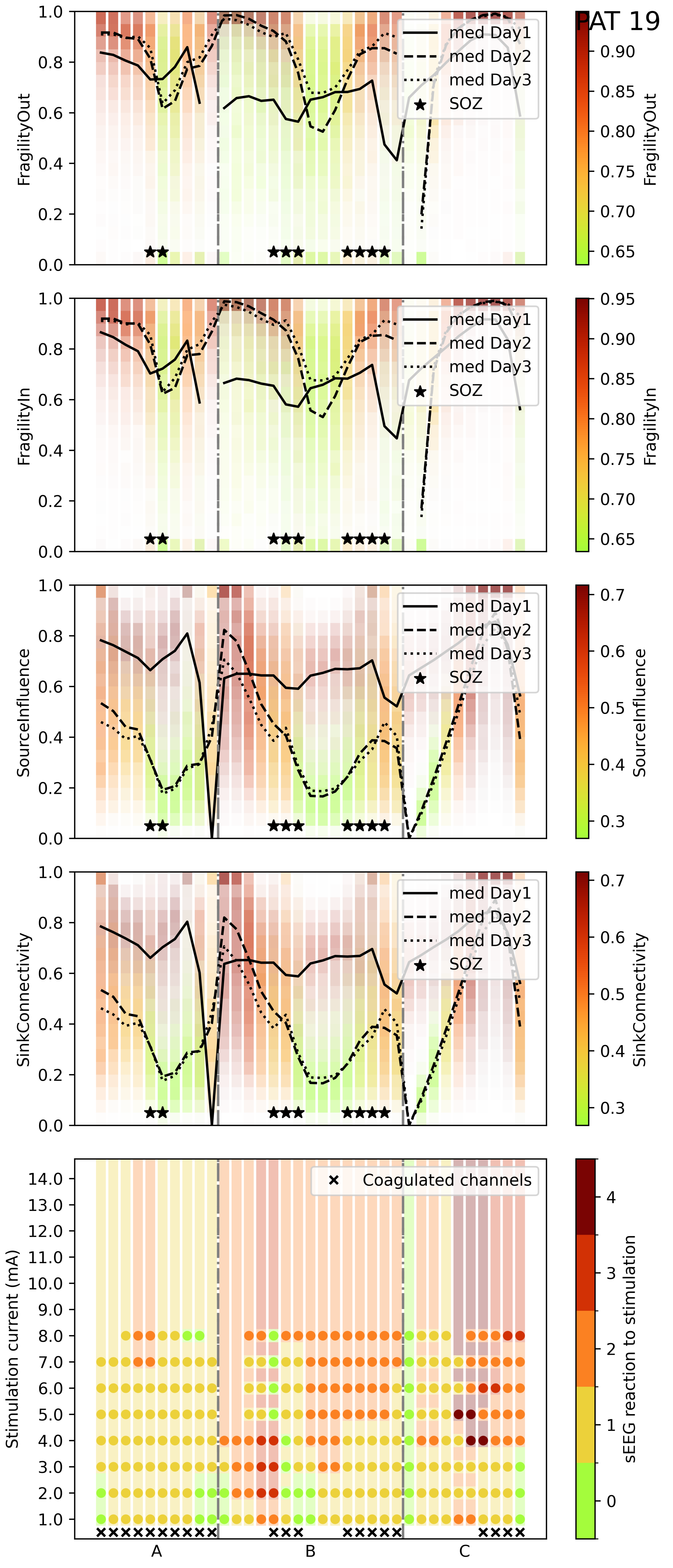

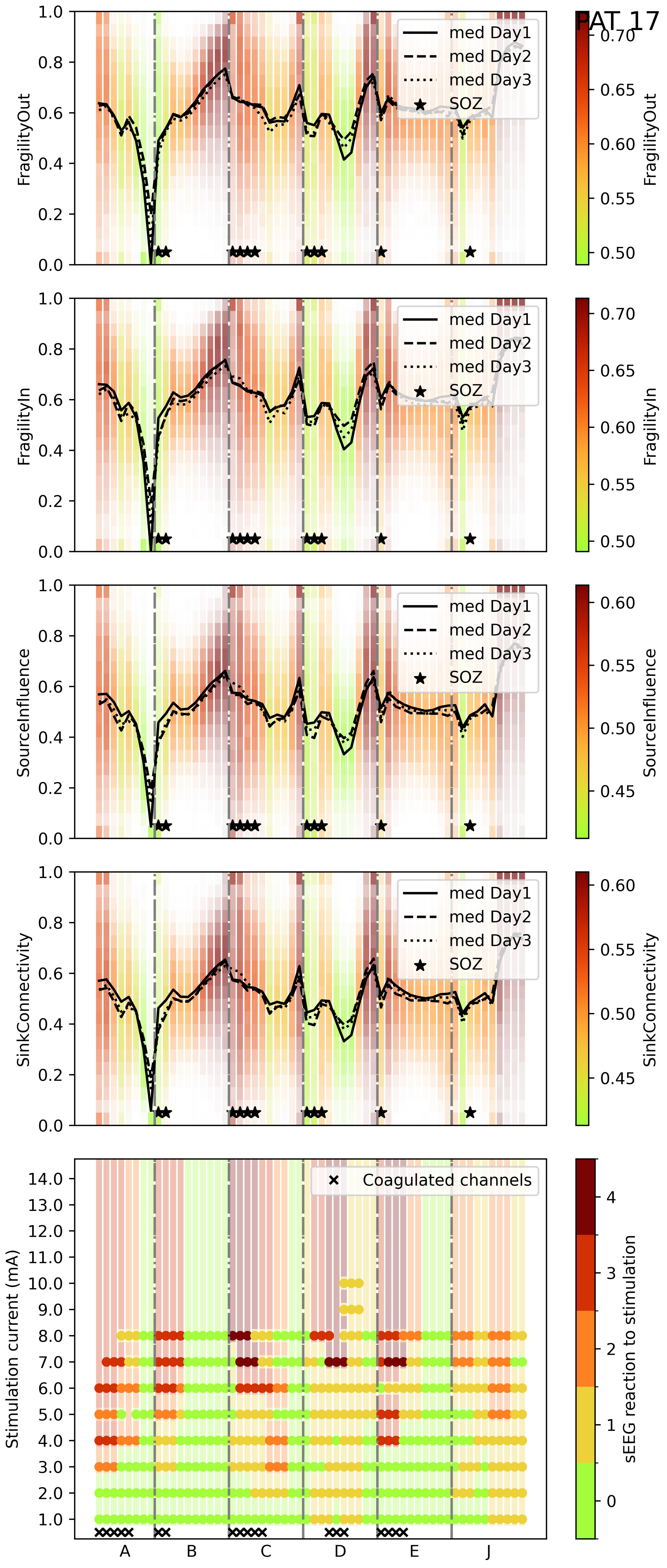

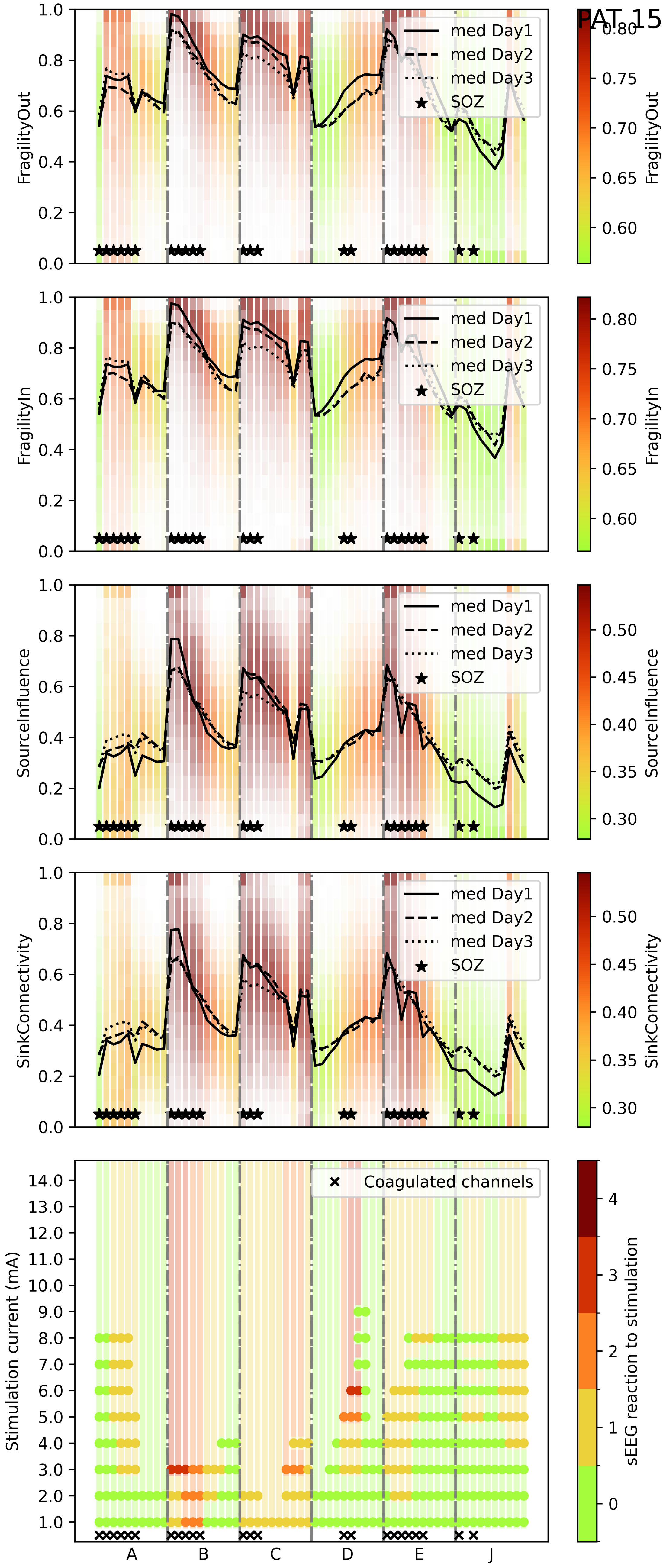

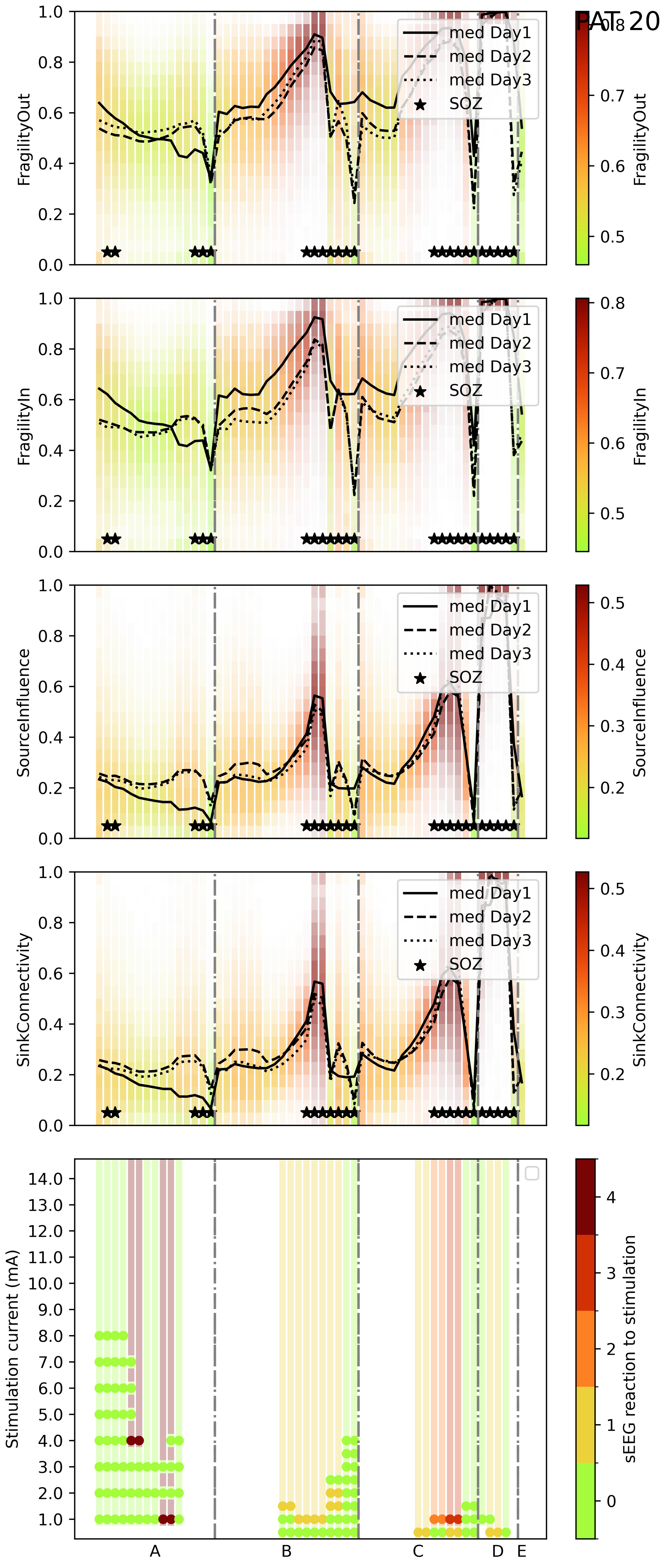
